# Data-driven clustering of mental health symptoms and brain functional connectivity signatures in transdiagnostic psychiatric inpatients

**DOI:** 10.1101/2024.12.30.24319777

**Authors:** Athina R. Aruldass, Shafi Rubbani, Peter Zhukovsky, Jennifer T. Sneider, Julia Cohen-Gilbert, Abdirahman Osman, Jocelyn Tatham, Joann Chen, Allyna-London Howell, Carmen Irujo, Lilikoi Bronson, Annah Rossvall, Lucie Duffy, Samantha Wong, Savannah Layfield, Fernando Rodriguez-Villa, Steven Gelda, Eliot Gelwan, Michael Leslie, Jane Eisen, Joseph Coletti, Diego A. Pizzagalli, Agustin G. Yip, Kerry J. Ressler, Nikolaos P. Daskalakis

**Author notes:** Corresponding author, Address correspondence to Dr Athina Aruldass: Neurogenomics & Translational Bioinformatics Laboratory (NG-TBL), Mclean’s Advanced Bioinformatics & Computational Discovery (ABCD) Hub, 1010 Pleasant Street, Belmont, MA 02478, USA. These authors contributed equally (Athina R. Aruldass, Shafi Rubbani).

## Abstract

Psychiatric disorders are notoriously heterogenous, often rendering diagnostic efforts challenging, and leading to poor therapeutic outcomes. The growing emphasis on ‘transdiagnostic’ approaches in psychiatry aligns well with the National Institute of Health-devised Research Domain Criteria (RDoc) framework that seeks to enable precision psychiatry. Here, we sought to identify transdiagnostic subgroups, which share behavioral and neural abnormalities, in a large population of psychiatric inpatients treated at a tertiary psychiatric hospital. The sample included 1,571 patients across five units within the Depression & Anxiety Disorders Division at McLean Hospital who completed self-assessments as part of an ongoing quality-of-care improvement project. Self-assessments included validated measures of depression, anxiety, anhedonia, trauma exposure, personality traits and substance misuse as well as broader screening questions. First, we identified naturally occurring transdiagnostic subgroups based on the self-reported questionnaires using consensus clustering to implement partition-around-medoids. In a subsample, we ascertained brain functional connectivity differences between the resultant subgroups using a multi-granular approach, i.e. whole-brain to connection-wise. In a *k*=2 partitioning solution, the first transdiagnostic cluster; C1 (N=809) had consistently higher means across all questionnaires compared to the second cluster; C2 (N=762). QIDS total score (P<0.0001, effect size = 0.54) and BASIS-24 derived total score (P<0.0001, effect size = 0.53) showed the largest difference between the groups. In follow-up imaging analysis (N=26) functional connectivity (FC) differences were observed connection-wise (P<0.0001) and between functional networks (P<0.0001), with C1 showing stronger FC than C2. Sex differences by cluster analyses revealed that females had higher BASIS-24 derived depression/functioning (C1: P<0.0001; C2: P<0.0001), and QIDS (C1|C2; P<0.0001 | P<0.0001) scores compared to males in both clusters. This was coupled with greater intermodular FC in males than females in C1 (P<0.0001) and C2 (P<0.0005). These results suggest that inpatients with transdiagnostic symptoms show biobehavioural abnormalities, underpinned by sex differences. Behaviourally, this is a function of acuity, i.e., severity of psychopathological symptoms, and not diagnosis. Biologically, the dysfunction captured here may span across various brain networks, rather than a singular region.

## INTRODUCTION

### Psychiatric diagnostics beyond the DSM

Classification of mental health disorders is traditionally represented by the Internal Classification of Disease (ICD) or Diagnostic & Statistical Manual of Mental Disorders (DSM), where often dichotomous categorical diagnoses are made following evaluation of a series of clinical symptoms. While nosology is needed for a variety of purposes in clinical medicine, since its inception and continual instrumentalization, psychiatric taxonomy has been a persistent subject of contention (1). Key issues pertain to (i) clinical heterogeneity, including shared symptomatology, that is, symptom profile across different diagnoses may overlap, and conversely, different symptom patterns resulting in the same diagnosis, as well as (ii) psychiatric comorbidity – co-occurrence of mental health disorders in an individual at a given time (1, 2). Adding to this complexity, diagnostic heterogeneity and comorbidity in psychiatry unequivocally result in further heterogeneity in disease trajectories, and treatment outcomes (3). Another problem with a categorical approach to psychopathological diagnosis is that of subsyndromal or subthreshold symptoms (4). Symptoms of a disorder usually present at a variety of levels of severity. However, if symptoms fall below a diagnostic threshold, diagnosis may be made without accounting for these symptoms, whose treatment could alleviate dysfunction and improve clinical condition. Is there a way then, to both minimize challenges associated with clinical heterogeneity and implement a more robust taxonomy system in place of the ICD/DSM?

### Transdiagnostic paradox in psychiatry: the ‘common factor’ vs ‘subtypes’

There is mounting support for a transdiagnostic approach in psychiatry (5), which cuts across existing traditional diagnostic boundaries or, more radically, sets them aside altogether, to provide novel insights into mental health difficulties. Evidence for this concept is the general psychopathology (*p*) factor (6) that is purported to capture latent ‘shared variation’ – hence, also known as ‘common factor’ – in behavioural symptomatology of internalizing (e.g. MDD, GAD), externalizing (e.g. ADHD, conduct disorder) and thought (e.g. schizophrenia, bipolar) disorders. Although ‘p’ remains elusive, prior hypotheses regarding key sources of latent shared variability include not only general psychopathology, as framed by the original authors (7), but also phenotypic factors such as psychosocial risk factors (8). A growing number of studies are also beginning to explore analogous factors to identify transdiagnostic biomarkers in psychiatry (9), such as genetic/heritable components (10–12) and neural substrates (13, 14).

This line of thinking on ‘non-p’ factors also complement the emerging ‘subtyping’ investigations in psychiatry seeking to identify nuances within a single disorder and across multiple disorders, via clustering methods (15, 16). The rationale is that resulting ‘subtypes’ or ‘subgroups’ might reveal latent properties distinguishing the subgroups as opposed to common latent feature(s) that are more common, e.g., transdiagnostic ‘p’ factor.

### Current investigation

In light of this, the present study aims to concurrently examine biobehavioral differences as well as similarities in transdiagnostic subgroups, specifically in an acute inpatient psychiatric setting. This was instrumentalized via (i) clustering of mental health and lifestyle instruments in a population of patients receiving psychiatric hospital level-of-care, and subsequently (ii) examining brain functional connectivity properties in resultant subgroups (**Figure 1**). Previous transdiagnostic clustering studies support the existence of diagnostically mixed subtypes across two or more disorders (17, 18). To our knowledge, Pelin et al. (2021) (18) constitutes the largest (N_discovery_=1,250) published transdiagnostic subtyping study. Here we expand on this with a larger sample size with likely more acute levels of symptoms. We hypothesized that resultant patient clusters or transdiagnostic subtypes would principally be discriminated by variations in severity of mental health symptoms, akin to majority of findings in single-disorder subtyping studies (19). We further hypothesized that the clinically-derived clusters would show related differences in brain functional connectivity.

**Figure 1:**
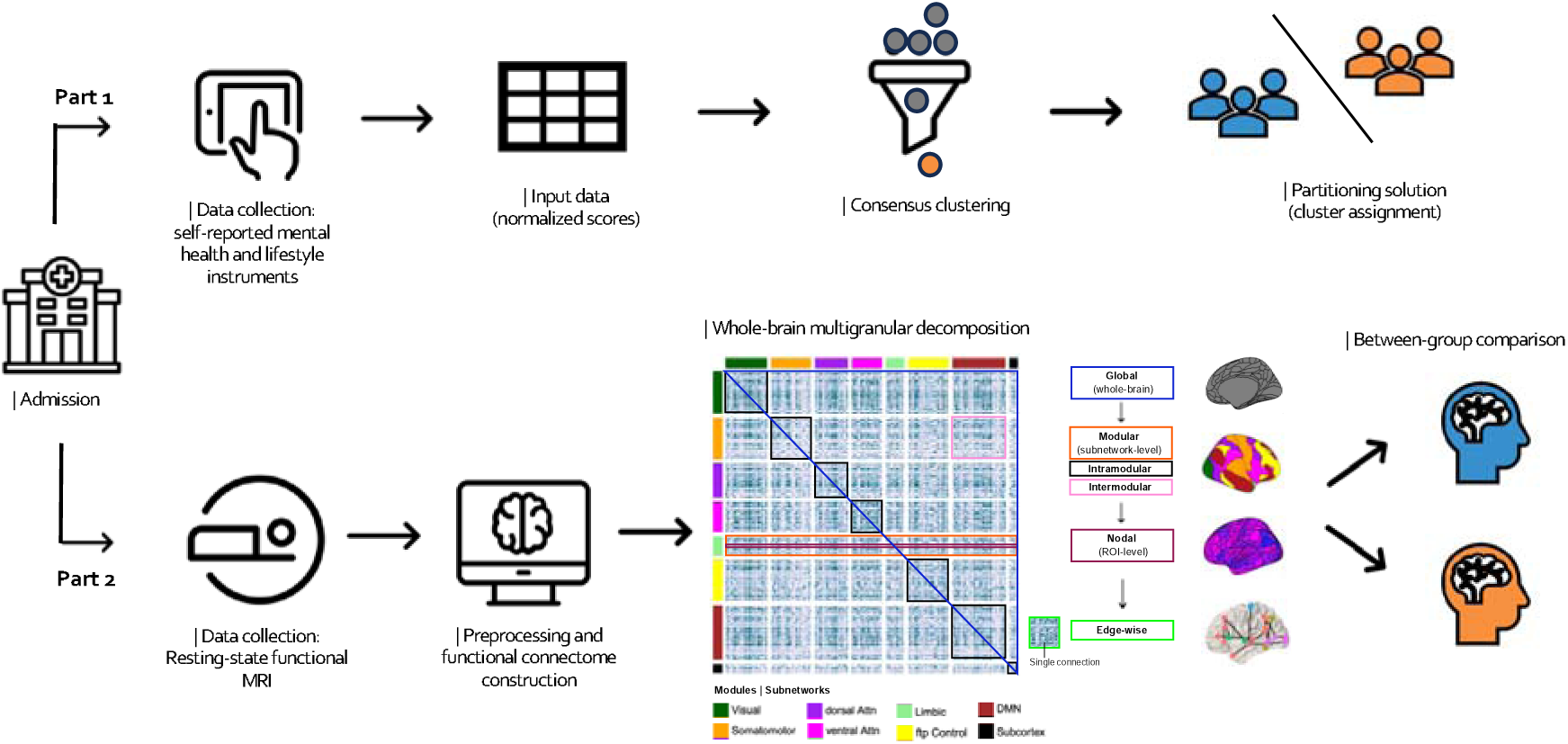
Study design and analyses overview. Part 1 of the study focused on unsupervised learning, that is, clustering of behavioral instruments ascertaining mental health symptoms and substance use. Part 2 of the investigation focused on examining brain-wide functional connectivity (FC) differences between resultant transdiagnostic subtypes. This was implemented through a multi-granular approach i.e., beginning with whole-brain global FC, intermodular FC (between functional networks) and intramodular FC (within functional networks), nodal or region-wise FC, and finally edge-or connection-wise FC.

## METHODS & MATERIALS

### Study design and Participants

Participants of the investigation were patients across five units of the Division of Depression & Anxiety Disorders at McLean Hospital. All procedures were approved by the Mass General Brigham Institutional Review Board (IRB), and all participants provided written informed consent. Between January 2018 and November 2023, research assistants approached patients and administered computer-based questionnaires (see Methods: Self-reported symptom scales) within 48 hours of admission as part of the Clinical Measurement Initiative (CMI; IRB# 2021P002444) (20), an ongoing patient care quality improvement/quality control (QI/QC) project. The self-assessments were administered on Apple iPads using Research Electronic Database Capture, *REDCap*. There were few exclusion criteria: high acuity preventing involvement, cognitive deficits limiting comprehension, and language barrier preventing communication.

### Self-reported symptom scales

All participants completed the following self-report standardized instruments assessing psychological and mental health symptoms (see Supplementary Materials Section S2): 24-item Behavior and Symptom Identification Scale (BASIS-24) (21), Quick Inventory of Depression Severity Scale (QIDS) (22), 7-item Generalized Anxiety Disorder (GAD-7) (23), Snaith-Hamilton Anhedonia Pleasure Scale (SHAPS) (24), PTSD checklist for DSM-V (PCL-5) (25), and the McLean Screening Instrument for Borderline Personality Disorder (MSI-BPD) (26). Participants also completed a series of questionnaires assessing substance use: Heaviness of Smoking Index (HSI; tobacco consumption) (27), concise version of the Alcohol Use Disorder Identification Test (AUDIT-C; alcohol consumption) (28), and the Drug Abuse Screening Test (DAST-10; drug use) (29). These questionnaires were completed after admission (i.e., within 48 hours). Participants completing the questionnaires beyond the indicated time frame were excluded. Clinical and sociodemographic information e.g. age, sex, personal information e.g. educational background, and family history were also collected as part of self-reported assessment. BASIS-24, QIDS, and GAD-7 drawn at discharge were used for validation.

### Consensus clustering

To identify naturally occurring transdiagnostic subgroups based on self-reported psychopathological symptoms and lifestyle habits, we performed consensus clustering (30) implemented via the *ConsensusClusterPlus* (v1.66.0) software (31) in R (v4.0.2). Consensus clustering improves robustness of clustering solution(s) by running multiple iterations of a given clustering algorithm through subsampling and evaluating stability of clustering assignments for each observation. The algorithm we implemented with consensus clustering was partitions around medoids (PAM). PAM, a *k*-medoids based algorithm, is a partitioning technique akin to the more commonly used *k*-means. However, where k-means solves the partitioning problem with respect to centroids (cluster means), PAM does so with respect to medoids (cluster centers being members of the cluster) that minimize distances or dissimilarities to all other observations within a cluster. The minimize error, PAM iterates over each cluster constituent as a medoid.

We used a subsampling parameter of 80% with 1000 iterations. The number of possible clusters (*k*) was selected to range from 2 to 10 to avoid excessive numbers of clusters that would not be meaningful. The ideal number of clusters were ascertained by evaluating the consensus matrix heatmap that visualize within-cluster (*k*) consensus score, and cumulative distribution function (CDF). The within-cluster consensus score (range; 0-1) is defined as the mean consensus value for all pairs of individuals belonging to the same cluster across iterations, i.e., at each iteration, if an individual or observation is assigned to the identical as previous iteration, value of 1 is ascribed, otherwise 0. We applied a strict threshold in defining ambiguous clustering, that is, a consensus score greater than 0 or less than 1 (as opposed to a more lenient boundaries e.g., 0.1 vs 0.9) was considered ambiguously clustered. A final value of 1 indicates strongest cluster stability.

### Partial least squares – discriminant analysis

To examine feature importance in cluster assignment, we performed partial least squares – discriminant analysis (PLS-DA), a supervised machine learning technique that is commonly used in multivariate analysis for dimensionality reduction, classification and feature selection (32). PLS-DA could be conceptualized as a ‘supervised’ version of Principal Component Analysis (PCA) and a categorical version of PLS-regression, in that it performs dimensionality reduction through linear transformation, but with full awareness of the class labels. It ascertains a linear relationship between two input matrices i.e., qualitative response variable or Y-variate (in our case, resulting cluster labels for each participant), and a set of continuous predictors or X-variate (in our study, 15 questionnaire-derived variables used in clustering analysis). Next, PLS-DA constructs a set of orthogonal components (akin to PCA components) that maximize the sample covariance between the response (Y) and the linear combination of the predictor variables (X). We implemented PLS-DA through *mixOmics* v6.26.0, in R v4.0.2 (33) with *k*=2 number of components, maximum 1000 number of iterations until convergence, convergence value *tol=1e-06*, and normalization applied to X matrix (scale parameter set to true; each data column is transformed to zero mean and unit variance). No parameter tuning, or cross-validation was performed as PLS-DA was purely implemented in our analyses to examine feature importance in clustering, and not for classifier training purposes.

### Resting-state functional MRI acquisition and preprocessing

A subset of CMI participants were also enrolled in the Psychometrics in McLean (IRB# 2016P001255) study. Data for Part 2 of our analysis were drawn from the Psychometrics at McLean MRI collection. Resting-state fMRI (rs-fMRI) data were acquired on a 3.0T Siemens Magnetom Prisma (Erlangen, Germany) scanner using multi-echo (ME) echoplanar imaging (EPI) sequence with: relaxation time (TR) = 1.33s; echo times (TE1, 2, 3, 4) = 13ms, 29ms, 48ms and 62ms; flip angle = 67 degrees; acquisition time = 7mins 40s = 300 volumes in each fMRI time series. ME-EPI data were collected as 60 slices (transversal) at 0 degrees to the A–P line with field of view = 216mm, for voxel resolution of 2.5 × 2.5 × 2.5mm. The first three volumes were discarded (dummy scans) and the remaining data were preprocessed. Standard preprocessing was performed using *fMRIPrep* (v23.0.2) with default parameters (34). As part of denoising, a 3mm FWHM kernel was applied for smoothing, and 24 nuisance parameters were regressed out (24 regressors; 6 motion derivatives (3 rotation and 3 translation), white Matter, CSF, and the respective first and second-order derivatives). Volume-to-surface mapping was then performed on the preprocessed and cleaned data. rs-fMRI data (volumetric standard space; MNI152NLin6Asym) were registered to standard space (surface; fsaverage HCP-MMP 1.0). Timeseries were extracted across 180 bilateral cortical regions (35) and 5 subcortical region-of-interests (ROIs) (FSL Harvard-Oxford atlas; accumbens, caudate, putamen, amygdala, hippocampus), resulting in 370 ROIs in total.

### Functional connectivity matrix construction and multi-granular connectome decomposition

*FSLNets* (https://fsl.fmrib.ox.ac.uk/fsl/fslwiki/FSLNets) was implemented via MATLAB to generate 370 x 370 symmetric connectivity matrices for each participant using the resultant timeseries. Full pair-wise correlations (normalized covariance) with Fisher r-to-Z transformation were estimated using the nets *nets_mats* function (‘corr’). Participants with high degree of head motion estimated by framewise displacement (FDrms > 0.3mm), were excluded (N = 5) (**Figure S2**). To gain more functional resolution of connectivity matrices, each ROI was also assigned to one of the 7 functional modules i.e. visual (V), somatomotor (SM), dorsal attention (DA), ventral attention (VA), limbic (L), frontoparietal control (FP), and default mode network (DMN), as described in Yeo et al. (2011), based on its co-localisation with these canonical resting-state networks. The remaining 10 subcortical regions were assigned to a subcortical (SC) module. Details of functional node-labelling schemes are further described in previous publications (36, 37). For statistical comparison, whole brain FC was decomposed using a multi-granular approach i.e., beginning with global FC, intermodular FC (between Yeo functional networks) and intramodular FC (within Yeo functional networks), nodal or region-wise FC (i.e., per ROI), and finally edge-or connection-wise FC **(Figure 1)**.

### Statistical methods

Correlation between clustering predictors were estimated using Spearman’s rank correlation. Between-group comparison to ascertain cluster differences were estimated using non-parametric Kruskal-Wallis test. Effect size reported were Cohen’s d estimated from η^2^ (eta squared) using the formula 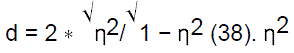 was initially derived from Kruskal-Wallis H-test using the formula 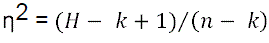 where *k* = number of groups, and *n* = number of observations. In Part 2, between-group comparison of brain FC were estimated using two-sample Kolmogorov-Smirnov tests and T-tests. P-values were adjusted for multiple comparisons with 5% false discovery rate (FDR). Post hoc estimation of interaction effect between factors was performed via two-way (2×2) ANOVA was performed. All statistical tests were performed in R (v4.0.2).

## RESULTS

### Sample characteristics

After quality control, N=1,571 participants remained for initial analyses (**Table 1**). The cohort was sex-balanced (46% ‘Male’), with a wide age range (18 – 81 years). Psychological instruments (BASIS-24, QIDS, SHAPS, GAD-7, PCL-5, MSI-BPD) were more strongly positively correlated with each other (ρ range: –0.077 – 0.815) than with demographic variables (ρ range: –0.198 – 0.169) and substance use instruments (ρ range: –0.167 – 0.516) (**Figure 2A**). There were significant negative correlations between demographic factors and symptom scales, including BASIS-24 derived depression functioning subscale (sex; P = 2.28e-15, ρ = –0.199), and total scores for QIDS (sex; P = 3.76e-14, ρ = –0.194) and MSI-BPD (age; P = 2.56e-14, ρ = –0.192). The correlation structure was retained under PTSD (PCL-5 total score) and sex-partialized correlation matrices. PTSD-adjustment resulted in weaker positive correlations (ρ range: –0.223 – 0.715) across all instruments, as well as the emergence of weak negative correlations between anhedonia (SHAPS) and substance use particularly (ρ range: –0.157 – –0.113), suggesting the influence of PTSD/trauma experience in substance abuse in the context of anhedonia (**Figure S3A**). In contrast, sex-adjustment enhanced positive associations between psychological instruments (ρ range: –0.184 – 0.807) (**Figure S3B**) and yielded strong positive correlations between age and mental health symptoms (ρ range: 0.584 – 0.651) that were not observed prior (**Figure 2A**). Females were significantly younger (**Figure S6A**; μFemale = 31.9year, μMale = 35.5years; P = 4.74e-9) than males in our cohort.

**Figure 2:**
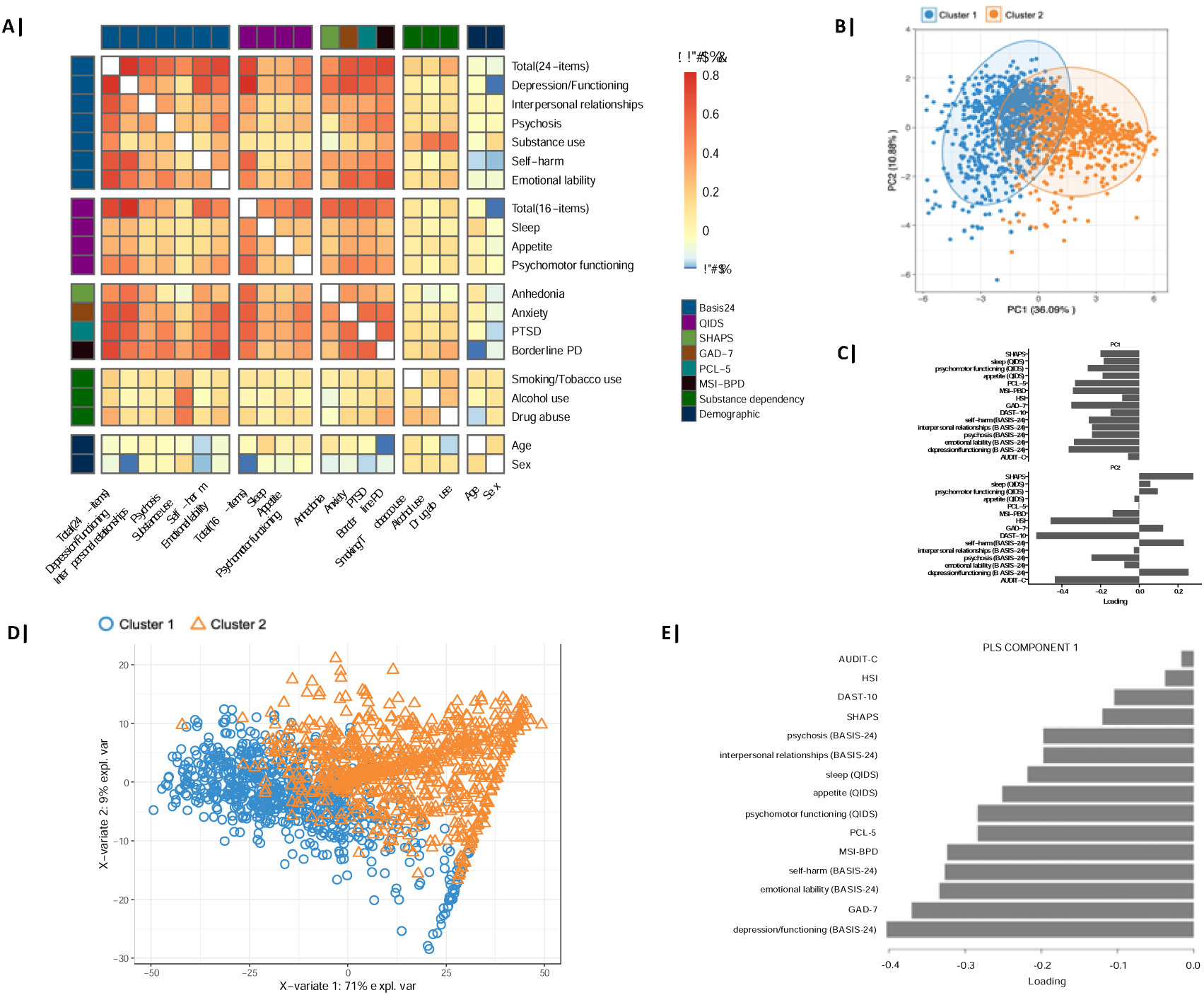
Association structure between clustering variables and clustering solution (k=2) and phenotypic predictors of transdiagnostic subgroups. (A) Behavioral instruments ascertaining mental health symptoms were generally more strongly correlated with each other, noted by the highly correlated (orange saturation) diagonal block. Of the demographic factors sex in particular was negatively correlated with select behavioral instruments/subscales suggesting that males (coded 1 in analyses) scored lower for these instruments/subscales compared to females. (B) Transdiagnostic subgroups show greater variation along PC1-axis, that is (C) a weighted-average of all input variables. The clusters showed significant difference in PC1 scores, with cluster 1 loading higher (more negative PC1 score) than cluster 2. correlation was estimated through Spearman’s method. (**D)** Partial least squares discriminant analysis (PLS-DA) for the predictors of Cluster 1 vs Cluster 2. The sample biplot depicts projection of study participants along first and second PLS components (x and y axes). Similar to our PCA projection, PLS component 1 appears to discriminate participants best, although discrimination is not clear or exclusive (overlap is notable). The plot also provides a premise on how novel samples would be classified based on position of along prediction background (blue and orange background) predicted given PLS-1 and PLS-2 scores. **(E)** PLS component 1 (X-variate) showed greatest covariance with Y-variate (71% explained variance) with BASIS-24 derived depression/functioning subscale, and anxiety loading most heavily in the component. These variables could thus be conceived as the most ‘important’ predictors of cluster labels for participants. Substance use dimensions in contrast, had marginal influence on clustering outcome.

**Table 1:** Sociodemographic and psychopathological variables in the analyzable cohort (N = 1,571). ^a^Behavior and Symptom Identification Scale; ^b^Quick Inventory of Depression Symptomatology; ^c^Generalized Anxiety Disorder (7-item); ^d^Snaith-Hamilton Pleasure Scale; ^e^PTSD Checklist for DSM-5; ^f^McLean Screening Instrument for Borderline Personality Disorder; ^g^ Heaviness of Smoking Index; ^h^ Alcohol Use Disorders Identification Index (Concise); ^i^Drug Abuse Screening Test (10-item); SD, standard deviation.

|  | Mean (SD) |
| --- | --- |
| **Demographics** |  |
| Total (N) | 1571 |
| Recruitment unit (N, %) |  |
| Unit 1 | 684 (43.5) |
| Unit 2 | 272 (17.3) |
| Unit 3 | 384 (24.4) |
| Unit 4 | 227 (14.4) |
| Unit 5 | 4 (0.2) |
| Sex (Male) (N, %) | 724 (46.1) |
| Race/Ethnicity (N,%) |  |
| White | 1173 (74.7) |
| Black | 113 (7.2) |
| Asian | 103 (6.6) |
| Age | 33.6 (14.1) |
| **Behavioral instruments** |  |
| BASIS-24<sup>a</sup> |  |
| Total | 40.0 (15.8) |
| Depression | 17.5 (6.7) |
| Psychosis | 1.9 (2.8) |
| Substance use | 2.9 (4.0) |
| Self-harm | 3.0 (2.4) |
| Emotional lability | 5.8 (3.2) |
| Relationships | 7.9 (4.5) |
| QIDS<sup>b</sup> |  |
| Total | 15.6 (5.8) |
| Sleep | 2.4 (0.8) |
| Appetite | 1.6 (1.1) |
| Psychomotor | 1.4 (1.0) |
| General Anxiety Disorder (GAD-7) | 13.4 (5.9) |
| Snaith-Hamilton Pleasure Scale (SHAPS) | 31.3 (8.3) |
| PTSD Checklist for DSM-5 (PCL-5) | 35.8 (22.1) |
| MSI-BPD<sup>c</sup> | 4.7 (2.8) |
| Heaviness of Smoking Index (HSI) | 0.4 (1.0) |
| Alcohol Use Disorders Identification Index (AUDIT-C) | 4.0 (2.9) |
| Drug Abuse Screening Test (DAST-10) | 1.6 (2.4) |

### Clustering outcome and optimal partitioning solution

Upon visual and empirical evaluation of consensus matrices (**Figure S4A**) and corresponding cumulative distribution function (CDF) plot (**Figure S4B**), *k=2* was selected as the best partitioning solution. The *k*=2 consensus matrix provided the best separation between clusters, with lower proportion of ambiguously clustered pairs (17.96%) across iterations, denser upper diagonal block (18.63%) and sparser off-diagonals compared to *k=3* (ambiguous clustering [‘0’] = 22.04%; consistent clustering [‘1’] = 2.6%) (**Figure S4A**). Similarly, inspection of CDF plot for *k=2* showed a sharp increase at consensus scores 0 and 1, with an evident horizontal line for intermediate consensus values, suggesting more stable clustering (**Figure S4B**).

### Cluster identity

To determine cluster identity, we examined differences in demographic factors, and input predictors between resultant clusters (cluster 1; C1 (N=809), cluster 2; C2 (N=762)). Both clusters showed significant differences across all behavioural instruments (Table S1). C1 had consistently higher mean values for QIDS total score (P<0.0001, Cohen’s d = 0.54), BASIS-24 total score (P<0.0001, Cohen’s d = 0.53), and BASIS-24 derived depression/functioning (P<0.0001, Cohen’s d = 0.52) compared to C2. Anxiety (GAD-7 total) (GAD-7 total score; P<0.0001, Cohen’s d = 0.42) and anhedonia (SHAPS total) were also significantly higher in C1 compared to C2 (P<0.0001, Cohen’s d = 0.31), although the effect sizes were smaller than questionnaires weighing on multiple psychopathologies. Substance use (drugs, alcohol and tobacco) was also significantly higher in C1 compared C2, although effect sizes were more minimal (P-values = P<0.0001 [DAST-10], 0.013 [AUDIT-C], 0.044 [HSI]; Cohen’s d range: 0.008 – 0.04) when contrasted against psychopathologies. There were a greater proportion of females in C1 than in C2 (C1|C2; N_females_ = 59.9% | 49.4%; P = 2.768e-05). To examine further the cluster separation, we performed principal component analysis (PCA) on clustering predictors (**Figure 2B-C**). PC1, a weighted average of all predictors was significantly different between the clusters (P<0.0001, t(1494)= –45.752; μC1 = –1.67 | μC2 = 1.77). PC2 also differed between the clusters, although the effect size of the difference was considerably smaller (P<0.0001, t(1522)= 5.973; μC1 = 0.179 | μC2 = –0.189) than PC1, evident from the limited cluster separation across the Y-axis in the biplot (**Figure 2B-C**).

Validation (N=1511) using identical subset of unobserved questionnaire data drawn at discharge supported the initial observations. Both clusters showed significant differences across all instruments apart from SHAPS (anhedonia), despite the effect sizes being smaller compared to admission-drawn measures (**Table S2**). C1 had consistently higher mean values, particularly for BASIS-24 total score (P<0.0001, Cohen’s d = 0.26), QIDS total score (P<0.0001, Cohen’s d = 0.25), and anxiety (GAD-7 total score; P<0.0001, Cohens’d = 0.53) compared to C2.

### Symptom scales that contributed most to separation of clusters

We used PLS-DA (**Figure 2D**) to find the weighted function of the 15 input variables (psychological instruments and substance use measures) that most accurately discriminated between C1 and C2. The first PLS component (X-variate 1) accounted for a large proportion (71.4%) of the variability in cluster assignments (**Figure 2D-E**). Consistent with our PCA results, BASIS-24 derived subscales (depression/functioning, emotional lability, self-harm), anxiety (GAD-7 total), and BPD (MSI-BPD total) were highly weighted on the first PLS component (–0.83, –0.36, –0.29 respectively; arbitrary unit), suggesting that a combination of these variables contributed most to C1/C2 assignment (**Figure 2E**). SHAPS and substance use instruments did not contribute greatly to clustering outcome as these were only more minimally different between clusters. SHAPS, HSI, DAST-10 and AUDIT-C also showed more limited variance in distribution within the cohort (**Figure S3C**).

### Brain functional connectivity differences

In Part 2 of our analyses, we examined brain FC differences between the C1 (N=25) and C2 (N=11). Examining distributions, relative to C2, C1 consistently had higher FC at global-level (**Figure 3A**; _μ_C1—C2 = 3.16—2.76; KS-statistic = 0.37, P=0.19) and edge-wise (**Figure 3B**; FC range: C1=-3.16–166.02, C2=-2.69–142.34; KS-statistic=0.13; P<0.0001). Across all intermodular connectivity, C1 generally had significantly higher FC than C2 (**Figure 3C**; FC range: C1=0.67–7.83, C2=0.43–8.22; KS-statistic =0.28; P<0.0001). Examination within specific module pairs showed nominal significance between the subcortex and three other networks: limbic (P_uncorrected_=0.043), default mode (P_uncorrected_=0.043) and fronto-temporo-parietal (P_uncorrected_=0.041), with mean connectivity being higher in C1 than C2 (**Tables S2-3**). At nodal-level, FC 33/360 cortical regions – including those within the dorsolateral prefrontal cortex (left p9-46v left area 46), anterior cingulate and middle prefrontal cortices (left p24), orbitofrontal cortex (left OFC), ventral visual stream (right VMV2), frontal and posterior opercula (left/right FOP1, right pFop), insula (left MI), somatosensory cortices (right area 3b,24dd) – and the left accumbens were higher in C1 than in C2 (**Figure 3D; Tables S3-4**). However, these differences did not survive FDR correction. Within functional modules (intramodular), the visual and ventral attentional networks had greatest differences between the clusters. However, these differences were not significant (**Figure 3E**).

**Figure 3:**
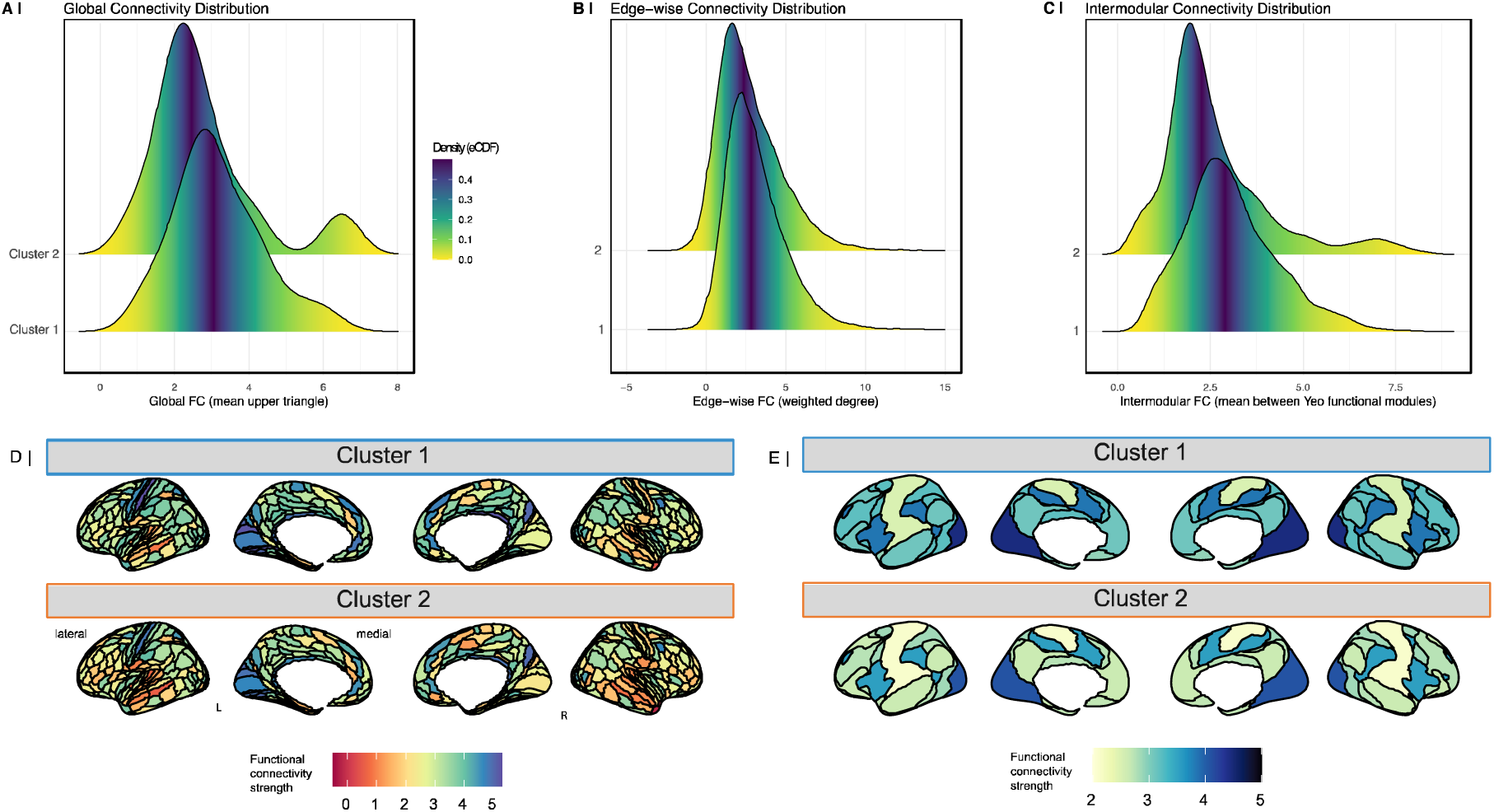
Brain functional connectivity differences in symptom-driven clusters. **(A)** Functional connectivity was not significantly different between clusters at global-level (estimated at individual-level). lHowever, significant difference in distribution was observed at **(B)** edge-wise (connection-wise; estimated from group-averaged connectivity matrices), and **(C)** at intermodular-level (between intrinsic functional networks; estimated at individual-level) with cluster 1 (C1) consistently showing stronger connectivity than cluster 2 (C2). **(D)** Few regions (nodes) showed nominal statistical significance. These include regions within the prefrontal cortices and opercular areas, with C1 showing stronger connectivity (more green/blue) than C2. **(E)** Intramodular (within functional networks) connectivity was not significantly different between clusters, although C1 again had generally higher connectivity (deeper blue intensities) that C2. statistical comparison was performed using Kolmogorov-Smirnov test.

### Sex-related behavioral and neurological differences within symptom clusters

As C1 had a significantly greater proportion of females than C2 (see Results ‘Cluster Identity’), we elected to further examine sex-related differences pre-clustering and within-cluster, in behavioural instruments most predictive of cluster assignments (BASIS-24 depression and GAD-7), as well as instrument with the greatest difference in terms of effect size (QIDS total score). Prior to clustering, females had significantly higher BASIS-24 derived depression/functioning (P<0.0001; _μ_F|_μ_M = 18.68639 | 16.09116), anxiety (GAD-7 total; P<0.0001; _μ_F|_μ_M = 13.92995 | 12.74217) and QIDS (total; P<0.0001; _μ_F|_μ_M = 16.53076 | 14.32249) scores than males (**Figure 4A-C** left panels). This observation persisted following clustering, where in both clusters, females similarly, had higher BASIS-24 derived depression/functioning (C1|C2; P<0.0001 | P<0.0001; _μ_F: 22.43277 | 13.85366; _μ_M: 21.19335 | 11.79389), and QIDS (C1|C2; P<0.0001 | P<0.0001; _μ_F: 19.88248 | 12.18560; _μ_M: 18.86 | 10.51) scores compared to males (**Figure 4A,C** right panels). Post-hoc analyses revealed that cluster-by-sex interaction effect was not significant for BASIS-24 depression [F(1, 1565) = 2.732, P=0.0986] and QIDS [F(1, 1526) =2.427 p=0.119], but was so for GAD-7 [F(1, 1565) =12.447, P=0.000431], given the opposite trend i.e., males higher than females, was noted within C1 (C1|C2; _μ_F: 16.87234 | 10.067039; _μ_M: 17.34688 | 8.884817). Nonetheless, this difference was not significant (C1|C2; P=0.066 | P<0.0001) (**Figure 4C** right panel). Validation using discharge data supported this sex-related observations, and further demonstrated consistency for GAD-7 as well. Before and after clustering, significant sex differences were observed for BASIS-24 depression/functioning (P<0.0001), QIDS total score (P<0.0001), and anxiety (GAD-7 total score; P<0.0001) (**Figure S5**).

**Figure 4:**
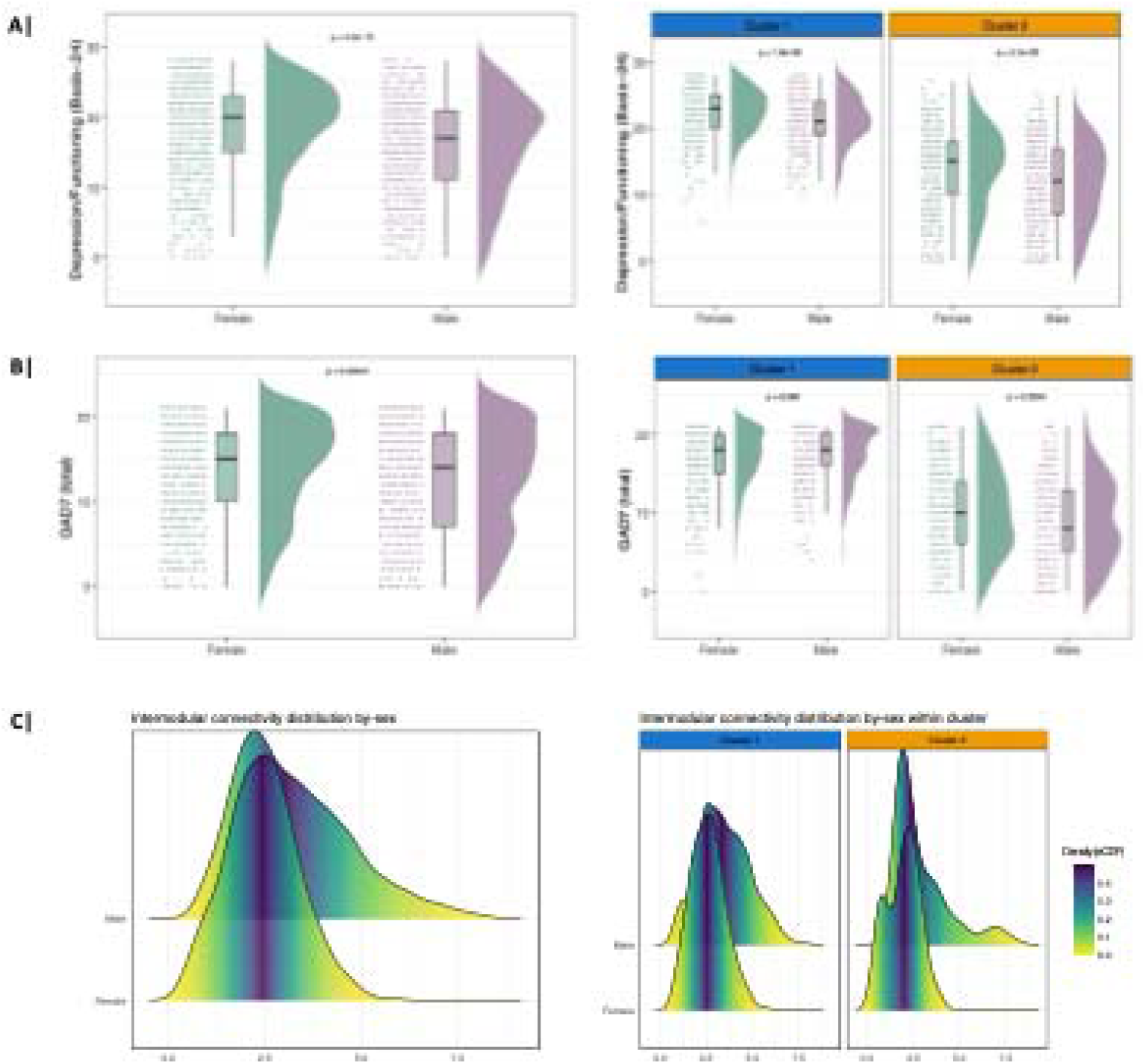
Sex-related differences did not drive symptom severity-related clusters. **(A-B)** Across behavioral instruments most predictive of clustering assignments, females consistently had higher scores suggesting greater severity endorsement than males, evident prior to clustering and within clusters. **(C)** By-sex intermodular-level (between intrinsic functional networks; estimated at individual-level) comparison before and after application of cluster assignments consistently showed statistically stronger connectivity in males compared to females. cluster 1 (Nmale | Nfemale = 13|12), cluster 2 (Nmale | Nfemale = 8|3). statistical comparison was performed using Kolmogorov-Smirnov test.

In relation to MRI findings, we elected to examine sex-related differences at intermodular-level upon observing the greatest between-cluster difference across intermodular FC distributions (see Results Brain functional connectivity differences). Overall, males had significantly higher intermodular FC compared to females (rangeM | rangeF = 0.837 – 8.218 | 0.434 – 5.723, D = 0.26701, p-value = 1.332e-15) (**Figure 4C** left panel). This observation was consistent within each cluster, in that males had significantly greater intermodular FC than females in C1 (rangeM | rangeF = 0.837 – 7.829 | 0.673 – 5.723; KS-statistic =0.31708; P<0.0001) and C2 (rangeM | rangeF = 0.963 – 8.218 | 0.434 – 3.821; KS-statistic =0.265; P=0.0003785) (**Figure 4C** right panel).

## DISCUSSION

In the current investigation, we present findings broadly supporting the transdiagnostic framework or dimensional approach to psychiatry. As hypothesized, our initial clustering analysis on self-reported mental health symptom and substance use questionnaires yielded patient clusters or transdiagnostic subtypes, that differed in severity of psychopathology, in particular depression and anxiety. Next, as predicted by our second hypothesis, we also demonstrated that the variation in acuity was coupled with brain functional connectivity differences between intrinsic resting-state networks (intermodular connections), spanning brain-wide connections.

### Single-diagnostic vs Transdiagnostic subtyping

Clustering and subtyping approaches are not new in psychiatry. Nonetheless, the majority of extant subtyping reports have typically focused on single-disorder subtyping, that is, discovery of subgroups within a single diagnostic category such as major depressive disorder (MDD), post-traumatic stress disorder (PTSD) and schizophrenia (17). While there is irrefutable value in such clustering efforts, Grisanzio *et al.* (2018) (17) accurately depicted a crucial gap that is not met through single-disorder subtyping – it ‘cannot address the need to characterize the heterogeneity and overlap of symptoms across diagnostic categories’. While psychiatric research has evolved over the past two decades exploring various transdiagnostic constructs/types e.g. within-spectrum (5), transdiagnostic subtyping that seek to classify patients beyond diagnostic criteria is arguably an important nascent research subdomain in psychiatry. As such, few studies have employed a comparable study design to the present investigation i.e. utilizing behavioral or mental health questionnaires as clustering variables. A previous study performed hierarchical clustering in a group of patients with primary diagnoses of either MDD, PTSD, and panic disorder, with comorbidity present (17). Analysis on self-reported negative mood, anxiety, and stress symptoms – ascertained through Depression, Anxiety and Stress Scale, version 21 (DASS-21) – yielded six transdiagnostic subgroups that differed in symptom profiles e.g. normative mood, anhedonia subgroups. This finding is unlike our outcome where resultant clusters were discriminated by acuity of symptoms, rather than psychopathological profiles. Another study [19] included risk factors, antecedent variables and concomitant indices that are associated with mental health disorders in adulthood such as general health, life events questionnaire (LEQ), childhood trauma questionnaire (CTQ) indices, maternal/paternal bonding and intelligence quotient (IQ). Using a discovery sample of N=1,250 (including N=590 health controls), the analysis yielded five clusters, where the first subgroup was mainly healthy controls, and remaining subgroups comprised distinct profiles of diagnoses, symptoms, and environmental risk factors. For the purpose of our analysis, we only included indices of mental health symptoms and related outcomes e.g. substance use, as opposed to risk factors. We designed our investigation in part to simplify interpretation of resultant transdiagnostic subgroups, and acknowledging the suggestion by Maj (2018) (39) who noted the need for a more detailed characterization of psychopathology of the individual case. Nonetheless, the author also emphasized the value of concurrent analyses unpacking vulnerability and protective factors, alluding to the exploration of antecedent variables such as family history, early environmental exposures, and the assessment of concomitant variables such as personality traits, socioeconomic status/resource, as well as cognitive functioning.

In relation to our Part 2 analyses on functional connectivity, while we note between-cluster differences, it is challenging to contextualize our findings with respect to wider research as extant studies on transdiagnostic subtyping have yet to characterize resultant clusters based on brain functional connectivity (17). Further, as discussed above, identity of our clusters was distinct from those in previous studies. It is worth noting however that currently, the weight of the literature on transdiagnostic subtyping stems from biotyping, that is, clustering of biological markers e.g. MRI markers (40–43), neurocognitive dysfunction (44). Such studies recognized that as with behavioral symptoms, there is also considerable overlap across diagnoses in regions showing abnormal connectivity, measured both functionally (i.e., temporal correlations between spatially disparate brain regions) and structurally (i.e., integrity of connecting white matter tracts of the brain) (42). A similar study focused on MDD utilized a more limited set of symptoms (Beck’s Depression Inventory-I; Beck’s Anxiety Inventory) for clustering (45) and reported associated frontotemporal network abnormalities between clusters that showed variations in symptom class. However, our current findings on intermodular and edge-wise differences perhaps conceptually align more with the cross-disorder brain dysconnectivity model that argues that ‘common symptoms arise from common circuit dysfunction’ (46). Indeed, emerging studies have begun reporting neural signatures that linked with transdiagnostic risk factors (13, 14, 47, 48). Thus, it is conceivable that behavioral symptom-driven clustering could reveal related brain functional network differences that cut across several diagnostic criteria that share psychopathologies.

### Sex-effects in self-reporting of mental health symptoms and implications on transdiagnostic psychiatry

Although the differences between clusters were greater, and duly the primary observation of our study, we deemed our sex-related findings were worthy of a discussion in the context of transdiagnostic and further, precision psychiatry. First, to emphasize, the analytic cohort was not confounded by sex-imbalance as the inpatient population was sufficiently balanced at 46.1% Male. We were additionally aware of potential recruitment unit biases e.g. existence of an all-female inpatient unit that could have disproportionately contributed to patient recruitment. However, male and female patients were represented across all five recruitment units within the initial analytic cohort, as well as within clusters (**Figure S6**). Within-cluster, females consistently scored significantly higher in instruments most predictive of the cluster assignments. Further, cluster-by-sex interaction effect was null, suggesting that sex differences observed in questionnaire outcomes is independent of cluster assignments. We venture that these observations could be attributable to sex-related healthcare/treatment disparities. ‘We have studies of fruit flies, mice, hamsters, frogs, monkeys, and men with this condition – but medical research using women as subjects just never occurred to anybody.’ – this caption from a 1991 editorial cartoon illustration (49) rather presciently introduced a key issue within healthcare research. Today, this has led to a healthcare system that is not attuned to female clinical presentations (50), heavily evidenced through studies on cardiovascular health (51–54) and pain management (55–57). In relation to psychiatric care, the 2021 Office for National Statistics (ONS) UK survey found that women struggled to attain ‘legitimacy’ for symptoms experienced during mental health crisis such as panic attacks or an episode of psychosis (58). Thus, it is conceivable that to attain equitable levels of care to males, females for example, would resort to presenting more severely at point-of-assessment (50, 59).

In relation our neuroimaging findings, we note that even following attrition attributed to unavailability of MRI scans, Male:Female composition between-clusters was comparable to Part 1 analyses in that C1 had more females, compared to C2 (C1; 50% males, C2;72.7% males). As such, on the one hand, observed differences in FC could be attributable to cross-symptom acuity/severity i.e. a transdiagnostic brain biomarker, or, on the other hand could be extrapolated to suggest sex-differences in functional connectivity patterns related to transdiagnostic symptoms. Neuroimaging related sex differences are arguably better-studied in the context of depression, including cerebral perfusion, structural and functional connectivity patterns (60). Some studies have also suggested that neuroimaging differences could be a product of sex and age interaction in mental disorders (61–63). We observed a significant difference in age between males and females in our cohort, and further within clusters (**Figure S6A**), with females being younger than males. However, these effect sizes were small, and we did not observe a main difference in age in non-sex-stratified cluster characterization (although age was marginally higher in C2). As such, we conclude that the observed FC differences were likely a product of transdiagnostic severity.

### Strengths & limitations

In light of the discussed findings, we noted key strengths of our study to be (i) use of varied and multiple self-reported instruments for psychopathology assessment, as well as (ii) a large, sex-balanced sample with a wide age range. Also noteworthy, we observed concordant findings to single-disorder data-driven clustering approaches i.e. the most common finding being total severity differences (19) that is yet to be demonstrated by previous transdiagnostic subtyping investigations. Therefore, our fundings argue that transdiagnostic subtypes are not simply a product of different symptom profiles, but also coupled with acuity of these symptoms. We also supported our cluster characterization with neuroimaging findings, although in a much more limited subset of participants. Taken together, we advocate for an interfacing between the RDoC and HiTOP framework, emphasizing both biobehavioral abnormalities and dimensional/spectral assessments of psychopathology (rather than binary per DSM). Taking heed from a report by Saunders *et al.* (2023) (64), we especially advocate for the integration/introduction of sex– and gender-specific surveys, diagnostic criteria, as well as treatment plans owing to the discrepancies in self-reporting of symptoms observed here. The typical handling of sex and gender in psychiatry research, and research at large has been as to consider these as ‘confounders’ and/or ‘biases’. In actuality, these are integral components of health and ill-health experience i.e. arguably, part of the disease/disorder that translate to related biological signatures.

An important limitation of our study is that we were unable to access clinical diagnoses i.e. primary diagnoses to confirm utility/accuracy of transdiagnostic status of our study cohort, though the majority of patients on the included units had primary diagnoses of MDD or anxiety related disorders. As all recruitment sites were represented in our sample, we interpreted the resultant cohort as transdiagnostic, comprising patients with both mood disorders, trauma and stressor-related disorders, and personality disorders (per DSM-V). We are at present unable to determine the proportion of these diagnoses within each cluster, although we note that this information would help to further understand and interpret the current findings. We also note that unlike previous investigations (17), our study design did not include a replication cohort. Nonetheless, we judiciously mitigated issues surrounding robustness and stability of clustering solution by implementing the clustering algorithm through consensus clustering with subsampling, as well as using unobserved discharge data to independently validate cluster identity. Our present investigation also did not include any healthy controls as the principal population were hospitalized psychiatric patients i.e. inpatients. Arguably, including healthy controls in the analysis might statistically confound clustering process. Future research could incorporate psychiatric outpatients as an intermediate comparison group, alongside health controls which aligns with an alternative approach to psychiatric diagnostics i.e. symptoms are a continuous function, and deviations could be represented through normative modelling (65). We also noted over our literature search on sex disparity in healthcare that the term ‘gender’ was frequently used in some early reports when ‘sex’ was meant. We were careful to not misrepresent findings from these studies, and further ensured that the authors clearly intended to examine sex differences despite using the word ‘gender’. Finally, we acknowledge that various clinical factors e.g. disease burden (duration of disorder, non-psychiatric comorbidities), admission history e.g. first hospitalization vs repeat, and sociodemographic variables e.g. race/ethnicity, educational background, socioeconomic status could equally moderate health experience, and therefore influence self-reporting of symptoms. This protected information was less available for our research use. However, consideration of these variables in future studies utilizing self-reported symptom scales is crucial.

## CONCLUSIONS

In conclusion, findings from this cross-sectional study suggest that heterogeneity within transdiagnostic inpatients was defined mostly by severity of mental health symptoms across various domains, and not by specific symptom profiles. Further, the neuroimaging observations point towards a putative transdiagnostic biomarker, supporting existing notions on ‘shared symptoms, share neurological dysfunctions’. The present results also suggest that these biobehavioral properties are linked with sex, a factor that should be examined more closely in context of transdiagnostic psychiatry.

## Data Availability

All data produced in the present study are available upon reasonable request to the authors.

## Acknowledgements

McLean Dissection of Anhedonia: From Neural Systems and Multiomics to Behavior”, awarded to DP, KR, and NPD, is supported by Wellcome Leap as part of the Multi-Channel Psych Program.

## Disclosures

Over the past 3 years, D.P. has received consulting fees from Boehringer Ingelheim, Compass Pathways, Engrail Therapeutics, Karla Therapeutics, Neumora Therapeutics (formerly BlackThorn Therapeutics), Neurocrine Biosciences, Neuroscience Software, Otsuka, Sage Therapeutics, Sama Therapeutics, Sunovion Therapeutics, and Takeda; he has received honoraria from the American Psychological Association, Psychonomic Society and Springer (for editorial work) and Alkermes; he has received research funding from the Bird Foundation, Brain and Behavior Research Foundation, Dana Foundation, Millennium Pharmaceuticals, NIMH, and Wellcome Leap; he has received stock options from Compass Pathways, Engrail Therapeutics, Neumora Therapeutics, and Neuroscience Software. K.J.R. has performed scientific consultation for Acer, Bionomics, and Jazz Pharma; serves on Scientific Advisory Boards for Sage, Boehringer Ingelheim, Senseye, Brain and Behavior Research Foundation and the Brain Research Foundation, and he has received sponsored research support from Alto Neuroscience. N.P.D. has been on scientific advisory boards for BioVie Inc., Circular Genomics, Inc., and Feel Therapeutics, Inc. for unrelated work. No funding from these entities was used to support the current work, and all views expressed are solely those of the authors. All other authors declare no competing interests.

The authors thank all the participants in the study and members of the Clinical Measurements Initiative (CMI), Psychometrics at McLean (PAM), and McLean1000 clinical and research teams – in particular, clinical staff, project coordinators, research assistants, medical imaging staff and laboratory staff.

## REFERENCES

1. Dalgleish T, Black M, Johnston D, et al.: Transdiagnostic Approaches to Mental Health Problems: Current Status and Future Directions. J Consult Clin Psychol 2020; 88:179– 195

2. Krueger RF, Eaton NR: Transdiagnostic factors of mental disorders. World Psychiatry 2015; 14:27–29

3. Plana-Ripoll O, Pedersen CB, Holtz Y, et al.: Exploring Comorbidity Within Mental Disorders Among a Danish National Population. JAMA Psychiatry 2019; 76:259–270

4. Bruno A, Iannuzzo F, Muscatello MRA: Comorbidity from a Categorical to a Transdiagnostic-Dimensional Approach: New Perspectives for Researchers and Clinicians. Clin Neuropsychiatry 2023; 20:7–8

5. Fusar-Poli P, Solmi M, Brondino N, et al.: Transdiagnostic psychiatry: a systematic review. World Psychiatry 2019; 18:192–207

6. Caspi A, Houts RM, Belsky DW, et al.: The p Factor: One General Psychopathology Factor in the Structure of Psychiatric Disorders? Clin Psychol Sci 2014; 2:119–137

7. Caspi A, Moffitt TE: All for One and One for All: Mental Disorders in One Dimension. AJP 2018; 175:831–844

8. Lahey BB, Krueger RF, Rathouz PJ, et al.: Validity and utility of the general factor of psychopathology. World Psychiatry 2017; 16:142–144

9. Sprooten E, Franke B, Greven CU: The P-factor and its genomic and neural equivalents: an integrated perspective. Mol Psychiatry 2022; 27:38–48

10. Selzam S, Coleman JRI, Caspi A, et al.: A polygenic p factor for major psychiatric disorders. Transl Psychiatry 2018; 8:1–9

11. Shi Y, Sprooten E, Mulders P, et al.: Multi-polygenic scores in psychiatry: From disorder specific to transdiagnostic perspectives. American Journal of Medical Genetics Part B: Neuropsychiatric Genetics 2024; 195:e32951

12. Grotzinger AD, Mallard TT, Akingbuwa WA, et al.: Genetic architecture of 11 major psychiatric disorders at biobehavioral, functional genomic and molecular genetic levels of analysis. Nat Genet 2022; 54:548–559

13. Romer AL, Knodt AR, Sison ML, et al.: Replicability of structural brain alterations associated with general psychopathology: evidence from a population-representative birth cohort. Mol Psychiatry 2021; 26:3839–3846

14. Romer AL, Elliott ML, Knodt AR, et al.: A pervasively thinner neocortex is a transdiagnostic feature of general psychopathology. Am J Psychiatry 2021; 178:174– 182

15. Feczko E, Miranda-Dominguez O, Marr M, et al.: The Heterogeneity Problem: Approaches to Identify Psychiatric Subtypes. Trends in Cognitive Sciences 2019; 23:584–601

16. Zhang W, Sweeney JA, Bishop JR, et al.: Biological subtyping of psychiatric syndromes as a pathway for advances in drug discovery and personalized medicine. Nat Mental Health 2023; 1:88–99

17. Grisanzio KA, Goldstein-Piekarski AN, Wang MY, et al.: Transdiagnostic Symptom Clusters and Associations With Brain, Behavior, and Daily Function in Mood, Anxiety, and Trauma Disorders. JAMA Psychiatry 2018; 75:201–209

18. Pelin H, Ising M, Stein F, et al.: Identification of transdiagnostic psychiatric disorder subtypes using unsupervised learning. Neuropsychopharmacol 2021; 46:1895–1905

19. Van Loo HM, De Jonge P, Romeijn J-W, et al.: Data-driven subtypes of major depressive disorder: a systematic review. BMC Med 2012; 10:156

20. Wong SA, Duffy LA, Layfield SD, et al.: Using electronic patient-reported measures to characterize symptoms and improvement in inpatient psychiatric units. Psychiatry Research 2022; 317:114839

21. Cameron IM, Cunningham L, Crawford JR, et al.: Psychometric properties of the BASIS-24© (Behaviour and Symptom Identification Scale-Revised) Mental Health Outcome Measure. Int J Psychiatry Clin Pract 2007; 11:36–43

22. Rush AJ, Trivedi MH, Ibrahim HM, et al.: The 16-Item Quick Inventory of Depressive Symptomatology (QIDS), clinician rating (QIDS-C), and self-report (QIDS-SR): a psychometric evaluation in patients with chronic major depression. Biol Psychiatry 2003; 54:573–583

23. Spitzer RL, Kroenke K, Williams JBW, et al.: A Brief Measure for Assessing Generalized Anxiety Disorder: The GAD-7. Archives of Internal Medicine 2006; 166:1092–1097

24. Snaith RP, Hamilton M, Morley S, et al.: A Scale for the Assessment of Hedonic Tone The Snaith-Hamilton Pleasure Scale 1995; 5

25. Blevins CA, Weathers FW, Davis MT, et al.: The Posttraumatic Stress Disorder Checklist for DSM-5 (PCL-5): Development and Initial Psychometric Evaluation. J Trauma Stress 2015; 28:489–498

26. Zanarini MC, Vujanovic AA, Parachini EA, et al.: A screening measure for BPD: the McLean Screening Instrument for Borderline Personality Disorder (MSI-BPD). J Pers Disord 2003; 17:568–573

27. Heatherton TF, Kozlowski LT, Frecker RC, et al.: Measuring the heaviness of smoking: using self-reported time to the first cigarette of the day and number of cigarettes smoked per day. Br J Addict 1989; 84:791–799

28. Bush K, Kivlahan DR, McDonell MB, et al.: The AUDIT Alcohol Consumption Questions (AUDIT-C): An Effective Brief Screening Test for Problem Drinking. Archives of Internal Medicine 1998; 158:1789–1795

29. Skinner HA: The drug abuse screening test. Addict Behav 1982; 7:363–371

30. Monti S, Tamayo P, Mesirov J, et al.: Consensus Clustering: A Resampling-Based Method for Class Discovery and Visualization of Gene Expression Microarray Data. Machine Learning 2003; 52:91–118

31. Wilkerson MD, Hayes DN: ConsensusClusterPlus: a class discovery tool with confidence assessments and item tracking. Bioinformatics 2010; 26:1572–1573

32. Ruiz-Perez D, Guan H, Madhivanan P, et al.: So you think you can PLS-DA? BMC Bioinformatics 2020; 21:2

33. Rohart F, Gautier B, Singh A, et al.: mixOmics: An R package for ‘omics feature selection and multiple data integration. PLoS Comput Biol 2017; 13:e1005752

34. Esteban O, Markiewicz CJ, Blair RW, et al.: fMRIPrep: a robust preprocessing pipeline for functional MRI. Nature Methods 2019; 16:111–116

35. Glasser MF, Coalson TS, Robinson EC, et al.: A multi-modal parcellation of human cerebral cortex. Nature 2016; 536:171–178

36. Váša F, Romero-Garcia R, Kitzbichler MG, et al.: Conservative and disruptive modes of adolescent change in brain functional connectivity. bioRxiv 2019; 604843

37. Aruldass AR, Kitzbichler MG, Morgan SE, et al.: Dysconnectivity of a brain functional network was associated with blood inflammatory markers in depression. Brain, Behavior, and Immunity 2021; 98:299–309

38. Fritz CO, Morris PE, Richler JJ: Effect size estimates: Current use, calculations, and interpretation. Journal of Experimental Psychology: General 2012; 141:2–18

39. Maj M: Why the clinical utility of diagnostic categories in psychiatry is intrinsically limited and how we can use new approaches to complement them. World Psychiatry 2018; 17:121–122

40. Goodkind M, Eickhoff SB, Oathes DJ, et al.: Identification of a Common Neurobiological Substrate for Mental Illness. JAMA Psychiatry 2015; 72:305–315

41. van den Heuvel MP, Sporns O: A cross-disorder connectome landscape of brain dysconnectivity. Nat Rev Neurosci 2019; 20:435–446

42. Vanes LD, Dolan RJ: Transdiagnostic neuroimaging markers of psychiatric risk: A narrative review. NeuroImage: Clinical 2021; 30:102634

43. Pfarr J-K, Meller T, Brosch K, et al.: Data-driven multivariate identification of gyrification patterns in a transdiagnostic patient cohort: A cluster analysis approach. Neuroimage 2023; 281:120349

44. Hilton RA, Tozzi L, Nesamoney S, et al.: Transdiagnostic neurocognitive dysfunction in children and adolescents with mental illness. Nat Mental Health 2024; 2:299–309

45. Maglanoc LA, Landrø NI, Jonassen R, et al.: Data-Driven Clustering Reveals a Link Between Symptoms and Functional Brain Connectivity in Depression. Biological Psychiatry: Cognitive Neuroscience and Neuroimaging 2019; 4:16–26

46. Buckholtz JW, Meyer-Lindenberg A: Psychopathology and the Human Connectome: Toward a Transdiagnostic Model of Risk For Mental Illness. Neuron 2012; 74:990–1004

47. Elliott ML, Romer A, Knodt AR, et al.: A Connectome-wide Functional Signature of Transdiagnostic Risk for Mental Illness. Biol Psychiatry 2018; 84:452–459

48. Alnæs D, Kaufmann T, Doan NT, et al.: Association of Heritable Cognitive Ability and Psychopathology With White Matter Properties in Children and Adolescents. JAMA Psychiatry 2018; 75:287–295

49. Hulme E: Women in research| Etta Hulme for Fort Worth Star Telegram [Internet]. Etta Hulme Cartoon Archive 1991; [cited 2024 Aug 12] Available from: https://libraries.uta.edu/ettahulme/browse/topics/research

50. Samulowitz A, Gremyr I, Eriksson E, et al.: “Brave Men” and “Emotional Women”: A Theory-Guided Literature Review on Gender Bias in Health Care and Gendered Norms towards Patients with Chronic Pain. Pain Res Manag 2018; 2018:6358624

51. Wenger NK: You’ve Come a Long Way, Baby*. Circulation 2004; 109:558–560

52. Wenger NK: Women and Coronary Heart Disease: A Century After Herrick. Circulation 2012; 126:604–611

53. Pencina MJ, D’Agostino RB, Larson MG, et al.: Predicting the 30-Year Risk of Cardiovascular Disease. Circulation 2009; 119:3078–3084

54. Vogel B, Acevedo M, Appelman Y, et al.: The Lancet women and cardiovascular disease Commission: reducing the global burden by 2030. The Lancet 2021; 397:2385–2438

55. Chen EH, Shofer FS, Dean AJ, et al.: Gender Disparity in Analgesic Treatment of Emergency Department Patients with Acute Abdominal Pain. Academic Emergency Medicine 2008; 15:414–418

56. Hoffmann DE, Tarzian AJ: The girl who cried pain: a bias against women in the treatment of pain. J Law Med Ethics 2001; 29:13–27

57. Mogil JS: Qualitative sex differences in pain processing: emerging evidence of a biased literature. Nat Rev Neurosci 2020; 21:353–365

58. Office for National Statistics: Office for National Statistics (ONS) Suicides in England and Wales: 2021 registrations. [Internet]. Suicides in England and Wales: 2021 registrations 2022; [cited 2024 Aug 12] Available from: https://www.ons.gov.uk/peoplepopulationandcommunity/birthsdeathsandmarriages/deaths/bulletins/suicidesintheunitedkingdom/2021registrations

59. Sharifian N, Kolaja CA, LeardMann CA, et al.: Racial, Ethnic, and Sex Disparities in Mental Health Among US Service Members and Veterans: Findings From the Millennium Cohort Study. American Journal of Epidemiology 2024; 193:500–515

60. Mohammadi S, Seyedmirzaei H, Salehi MA, et al.: Brain-based Sex Differences in Depression: A Systematic Review of Neuroimaging Studies. Brain Imaging Behav 2023; 17:541–569

61. Giedd JN, Raznahan A, Mills KL, et al.: Review: magnetic resonance imaging of male/female differences in human adolescent brain anatomy. Biol sex dif 2012; 3:19

62. Ruigrok ANV, Salimi-Khorshidi G, Lai M-C, et al.: A meta-analysis of sex differences in human brain structure. Neurosci Biobehav Rev 2014; 39:34–50

63. Raznahan A, Disteche CM: X-chromosome regulation and sex differences in brain anatomy. Neuroscience & Biobehavioral Reviews 2021; 120:28–47

64. Saunders EFH, Brady M, Mukherjee D, et al.: Gender differences in transdiagnostic domains and function of adults measured by DSM-5 assessment scales at the first clinical visit: a cohort study. BMC Psychiatry 2023; 23:709

65. Marquand AF, Rezek I, Buitelaar J, et al.: Understanding Heterogeneity in Clinical Cohorts Using Normative Models: Beyond Case-Control Studies. Biol Psychiatry 2016; 80:552–561

