## Supplementary_Materials for "Data-driven clustering of mental health symptoms and brain functional connectivity signatures in transdiagnostic psychiatric inpatients"

**Authors & Affiliations:**

Athina R. Aruldass∗†, Shafi Rubbani∗, Peter Zhukovsky, Jennifer T. Sneider, Julia Cohen-Gilbert, Abdirahman Osman, Jocelyn Tatham, Joann Chen, Allyna-London Howell, Carmen Irujo, Lilikoi Bronson, Annah Rossvall, Lucie Duffy, Samantha Wong, Savannah Layfield, Fernando Rodriguez-Villa, Steven Gelda, Eliot Gelwan, Michael Leslie, Jane Eisen, Joseph Coletti, Diego A. Pizzagalli, Agustin G. Yip, Kerry J. Ressler, Nikolaos P. Daskalakis.

From: Division of Depression & Anxiety Disorders, McLean Hospital (ARA, SR, AO, JT, JC^1^, ALH, CI, LB, AR, LD, SW, SL, FRV, SG, ML, JE, JC, DAP, AGY, KJR, NPD); Harvard Medical School (ARA, PZ, JTS, JCG, DAP, AGY, KJR, NPD); Stanley Center for Psychiatric Research, Broad Institute (ARA, NPD); Centre for Depression, Anxiety & Stress Research, McLean Hospital (PZ, DAP); McLean Imaging Center (JTS, JCG, DAP); Dissociative Disorders and Trauma Inpatient Program; McLean Hospital (EG).

∗ these authors contributed equally

Neurogenomics & Translational Bioinformatics Laboratory (NG-TBL), McLean Waverley Place, 1010 Pleasant Street, Belmont, MA 02478, USA.

**Contents**

*Supplementary Methods & Results*

| **Section S1** Participants | 3 |
| --- | --- |
| **Section S2** Psychological evaluation and behavioral assessment | 4 |
| **Section S3** Neuroimaging | 6 |
| **Section S4** Partial correlation and distributions of clustering input variables | 7 |
| **Section S5** Clustering solution & Between-cluster differences | 8 |
| **Section S6** Sex-related differences in self-reporting and age | 9 |
| **Section S7** Distributional differences in recruitment site: within- cohort and cluster | 10 |
| **Section S8** Between-cluster multi-granular functional connectivity differences | 11 |

*List of Figures*

| **S1** Participant recruitment flow-chart | 3 |
| --- | --- |
| **S2** Additional motion correction | 6 |
| **S3** Sex and trauma partialized correlation matrices | 7 |
| **S4** Optimal clustering solution | 8 |
| **S5** Sex differences within-cohort and within–cluster at discharge | 9 |
| **S6** Distributional differences in recruitment sites | 10 |

*List of Tables*

| **S1** Cluster characterization for *k*=*2* partitioning solution | 8 |
| --- | --- |
| **S2** k=2 Cluster identity validation using discharge data | 11 |
| **S3** Between-cluster global and modular functional connectivity |  |
| **S4** Between-cluster nodal functional connectivity | 12 |

***Supplementary Methods***

**Section S1 Participants**

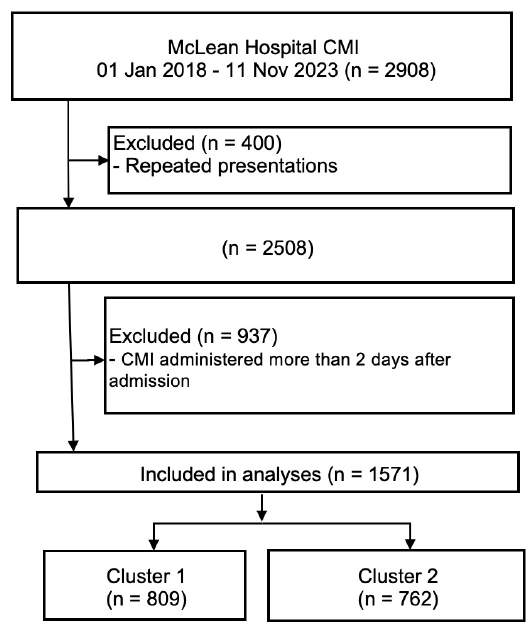

**Figure S1: Flow diagram outlining progression of participants through to analyses.**

**Section S2 Psychological evaluation and behavioral assessment**

All participants completed the following self-report standardized instruments: 24-item Behavior and Symptom Identification Scale (BASIS-24), Quick Inventory of Depression Severity Scale (QIDS), 7-item Generalized Anxiety Disorder (GAD-7), Snaith-Hamilton Anhedonia Pleasure Scale (SHAPS), PTSD checklist for DSM-V (PCL-5), McLean Screening Instrument for Border- line Personality Disorder (MSI-BPD), Heaviness of Smoking Index (HSI; tobacco consumption), concise version of the Alcohol Use Disorder Identification Test (AUDIT-C; alcohol consumption), and the Drug Abuse Screening Test (DAST-10; drug use). Clinical and sociodemographic information. age, sex, personal information e.g. educational background, and family history were also self-reported.

**Behavior and Symptom Identification Scale (BASIS-24)** (Cameron et al, 2007; Eisen et al., 2006). The BASIS-24 is a broad psychiatric symptom scale, assessing symptoms over the past week. The main 24 items weigh towards 6 subdomains: (i) Depression/Functioning (‘Feel sad or depressed?’, items #1–3,10), (ii) Interpersonal relationship (e.g. ‘Get along with people in your family?’, items #4–9), (iii) Self-harm (‘Think about ending your life?’; items #11,20), (iv) Emotional lability (‘Have mood swings?’, items #12,18–19), (v) Psychosis (e.g. ‘Hear voices or see things?’, items #13–17), and (vi) Substance abuse/dependence (e.g. ‘Did you have an urge to drink alcohol or take street drugs?’; items #21–24). Participants are asked to rate items on a 5-point Likert type rating scale from 0 (none of the time) to 4 (all of the time), with higher scores indicating worse functioning. Subscale scores range from 0–8 (self-harm) to 0–24 (de- pression/functioning) and total scores reflect overall functioning. The questionnaire also includes 12 demographic questions.

**Quick Inventory of Depression Severity Scale (QIDS) – Self-Reported (**Rush et al, 2003; Trivedi et al.). The QIDS-SR is a 16-item instruments to assess depressive symptom sever- ity. QIDS converts responses to 16 separate items into the nine DSM-IV symptom criterion domains for MDD. comprising (i) low mood; (ii) concentration; (iii) self-criticism; (iv) suici- dal ideation; (v) interest; (vi) energy/fatigue; (vii) sleep disturbance (initial, middle, and late insomnia or hypersomnia); (viii) decrease/increase in appetite/weight; and (ix) psychomotor ag- itation/retardation. Three of the subdomains are derived via subscales i.e. ‘sleep disturbance’ (items #1–4), (ii) ‘appetite changes’ (items #6–9), and (iii) ‘psychomotor impairments’ (items #15–16). The total score is calculated by summing scores across the subdomains and range from 0 to 27.

**McLean Screening Instrument for Borderline Personality Disorder (MSI-BPD)** (Zanarini, Vujanovic, Parachini, Boulanger, Frankenburg, & Hennen, 2003). The MSI-BPD is a brief 10- item measure screening for borderline personality disorder traits in a yes/no format. The instru- ment is based on a subset of the questions that comprise the borderline module of the Diagnostic Interview for DSM-IV Personality Disorders or DIPD-IV, and as such has excellent sensitivity as well as specificity for BPD diagnosis. The ten items of the MSI-BPD are written such that a positive response indicates the presence of BPD symptoms. Each item of this instrument is rated on a dichotomous scale with 1 corresponding to ’present’ and 0 corresponding to ’absent’. A score of 7 or more yields a diagnosis of BPD.

**Snaith-Hamilton Pleasure Scale (SHAPS)** (Snaith et al., 1995). This 14-item self-reported instrument was used to measure hedonic tone or more strictly, anhedonia – one of the core symp- toms of depression (Snaith et al., 1993). Each item has a 4-level Likert scale i.e. ‘Definitely agree’, ‘Agree’, ‘Disagree’, ‘Strongly disagree’. Per our scoring system, the former two ‘Agree’ responses receive a score of 4 and 3 respectively, whilst the latter two ‘Disagree’ responses re- ceive 2 and 1 respectively. Thus, total scores range from 14 to 56, with higher scores indicating higher levels of current anhedonia or reduced hedonic experience.

**7-item Generalized Anxiety Disorder (GAD-7)** (Spitzer et al., 2006). The GAD-7 consists of seven items measuring worry and anxiety symptoms. The seven items assess (i) feeling nervous, anxious, or on edge; (ii) being able to stop or control worrying; (iii) worrying too much about dif- ferent things; (iv) trouble relaxing; (v) being restless; (vi) becoming easily annoyed or irritable; and (vii) feeling afraid as if something awful might happen. Participants indicated the frequency with which they experienced specific symptoms over the past 24 hours on a 4-point Likert-type scale (from 0 = not at all to 3 = nearly all the time). Total scores range from 0 to 21, with higher scores reflecting greater anxiety severity. Scores above 10 are considered to be in the clinical range for GAD diagnosis.

**PTSD checklist for DSM-V (PCL-5)** (Weathers et al., 2013; Belvin et al., 2015). The PTSD Checklist (PCL) (Weathers, Litz, Herman, Huska, Keane, 1993) was one of the mostly widely used screening measures for PTSD (Elhai, Gray, Kashdan, Franklin, 2005; McDonald Calhoun, 2010). The PCL-5 is one of few PTSD self-report instruments that have been updated to re- flect DSM-5 changes to PTSD diagnostic criteria. The instrument includes 20 self-report items based on the DSM-5 symptoms of PTSD. Respondents report how much they were bothered by a symptom over the past month using a 5-point Likert scale (0 = ‘Not at all’; 1 = ‘A little bit’; 2 = ‘Moderately’; 3 = ‘Quite a bit’; 4 = ‘Extremely’). Total score can range from 0 to 80, with a cutoff score of 31-33 suggesting best diagnostic utility i.e. shows highest predictability of CAPS-5 (clinician rated PTSD assessment) diagnoses.

**Section S3 Neuroimaging**

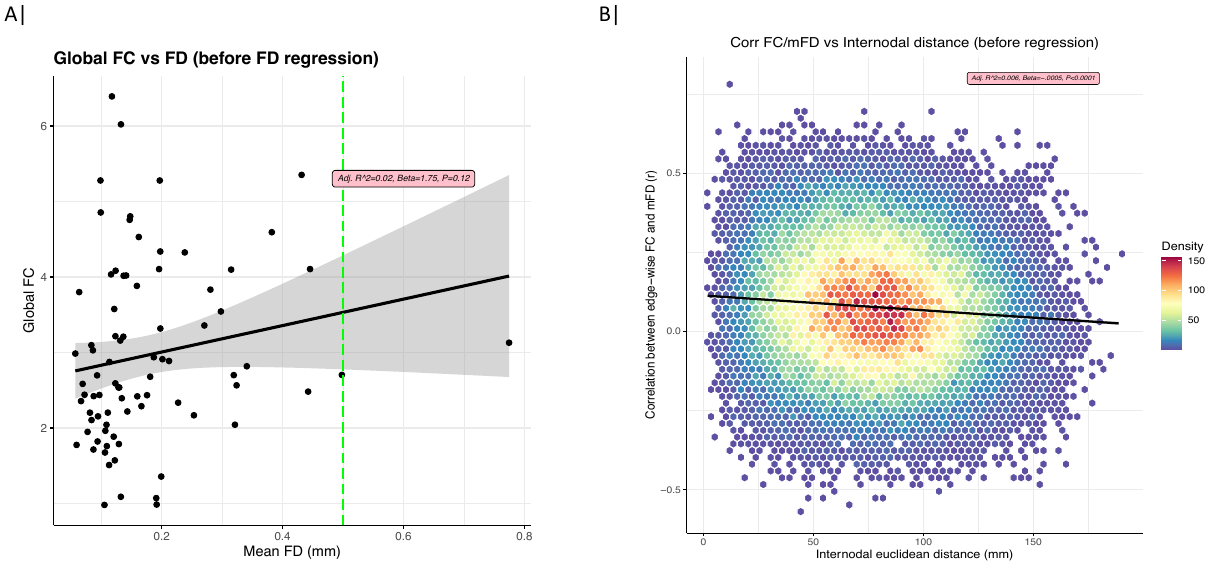

**Figure S2: Additional motion correction in FC matrices.** After standard preprocessing of fMRI timeseries, we critically took additional measures to ascertain any residual motion-related effects on functional connectome. Our diagnostic procedure comprised **(A)** assessing for effects of motion (indexed by root mean square framewise displacement; FDrms) on global FC, and **(B)** determining likelihood of edge-wise correlation between FC and mean FD, and if this association fluctuated with internodal distance† i.e. distance between two connecting nodes, as previously demonstrated (Power et al., 2012; Satterthwaite et al., 2016). From the diagnostic plots (Before) we noted **(A)** a global non-significant effect of motion on FC, in that global FC increased with motion (β=1.75, R^2^adj=0.02, P=0.12), and **(B)** on average edges showed correlation with mean FD (indicated by y-intercept = 0.13) and this correlation decreased marginally (negative slope; β=-0.0005) with increasing distance between nodes i.e. long-range edges were more vulnerable to motion-related effects compared to short-range edges (R^2^adj= 6x10^-3^, P< 0.0001). In addition to excluding participants with mean FD > 0.3mm, we performed edge-wise regression of FDrms to remedy the observed association between functional connectivity and motion.^†^distance spanning across nodes was defined as Euclidean distance; C[i,j] = sqrt[(∆*X*)2 +(∆*Y*)2 +(∆*Z*)2] ]; *where* ∆X=X*i* –X*j*, i=node1, j=node2; X, Y and Z = centroid coordinates in MNI space for each mapped parcel.*p < 0.05; **p < 0.01; ***p < 0.001

**Supplementary Results**

**Section S4 Partial correlation between input variables for clustering and distribution of input variables**

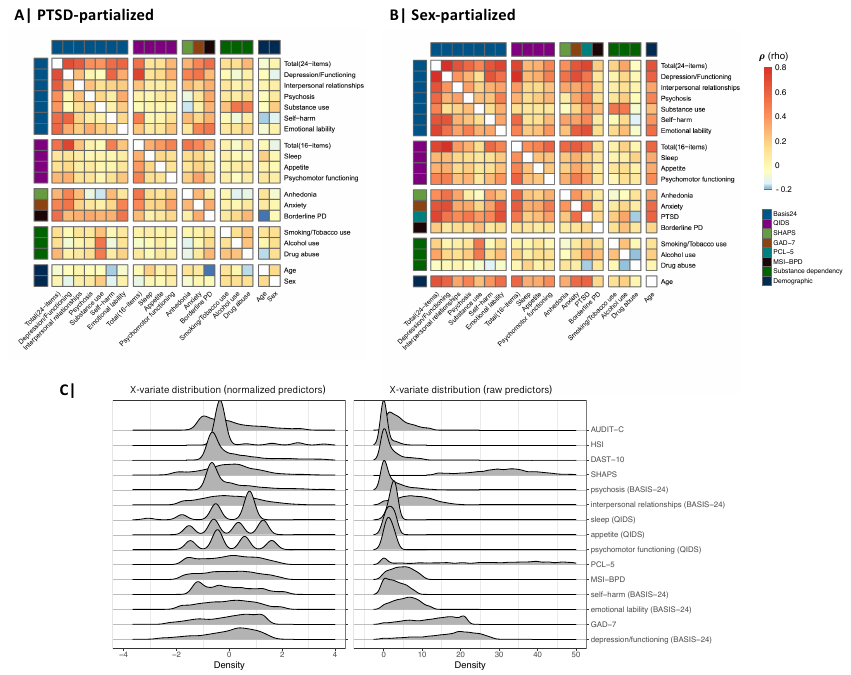

**Figure S3: Partialized correlation matrices.** **(A)** Correlation between clustering variables after adjusting for PTSD (PCL-5 total score). **(B)** Correlation between clustering variables after adjusting for sex. Correlation was estimated Spearman’s method. **(C)** Across input variables for clustering, BASIS-24 derived depression/functioning subscale, anxiety, BASIS-24 derived emotional lability subscale, BASIS-24 derived self-harm, borderline psychopathology and PTSD have greater dispersion i.e. hold greater variance, compared to substance use measures.

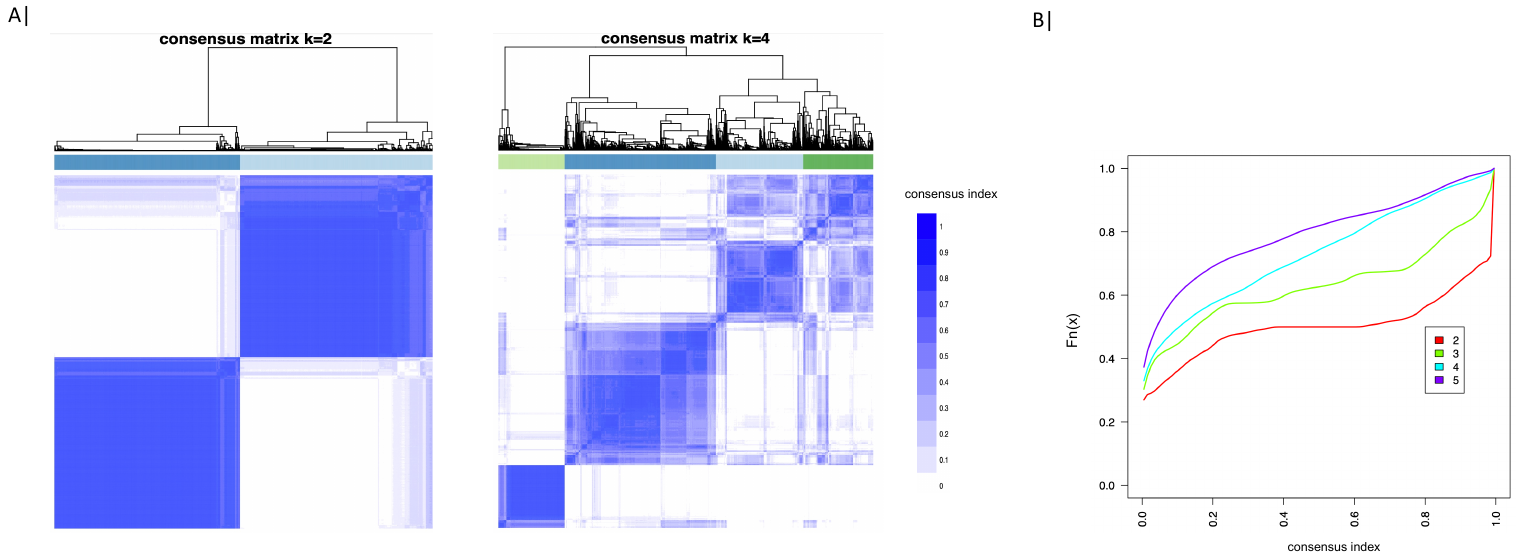
**Section S5 Clustering solution & Between-cluster differences**

**Figure S4: Optimal clustering solution.** **(A)** We applied a strict threshold in defining ambiguous clustering, that is, a consensus score greater than 0 or less than 1 (as opposed to a more lenient boundaries e.g., 0.1 vs 0.9) was considered ambiguously clustered. A final value of 1 indicates strongest cluster stability. As such, visually the optimal clustering solution would have the ‘cleanest’ consensus matrix with less off-diagonals (left panel) vs a poor clustering solution would be less ‘clean’ with more off-diagonals (right panel). **(B)** Correspondingly, the optimal clustering solution would have a higher frequency of ‘0’ and ‘0’, denoted by steeper increase in CDF trendline.

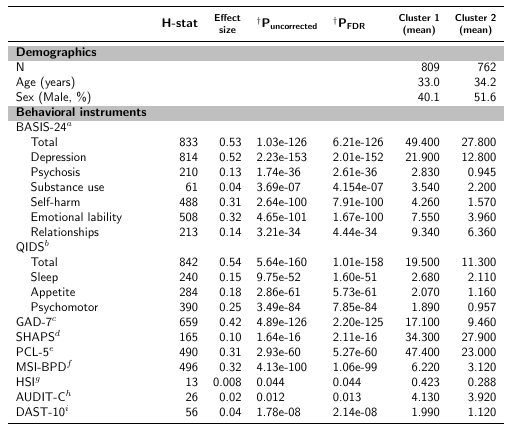

**Table S1: Behavioral and demographic characterization of resultant clusters (*k*=*2*).** aBehavior and Symptom Identification Scale; bQuick Inventory of Depression Symptomatology; cGeneralized Anxiety Disorder (7-item); dSnaith-Hamilton Pleasure Scale; ePTSD Checklist for DSM-5; fMcLean Screening Instrument for Borderline Personality Disorder; g Heaviness of Smoking Index; h Alcohol Use Disorders Identification Index (Concise); iDrug Abuse Screening Test (10-item); ^†^statistical comparison performed using Kruskal-Wallis test; ‡ Effect size reflect Cohen’s *d .*

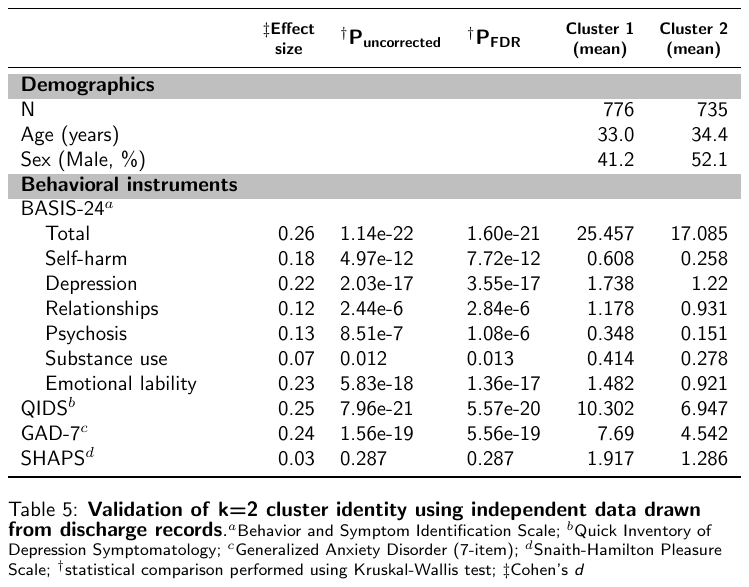

**Table S2: Validation of k=2. Cluster identity using independent data drawn from discharge records.** aBehavior and Symptom Identification Scale; bQuick Inventory of Depression Symptomatology; cGeneralized Anxiety Disorder (7-item); dSnaith-Hamilton Pleasure Scale; ^†^statistical comparison performed using Kruskal-Wallis test; ‡ Effect size reflect Cohen’s *d .*

**Section S6 Sex-related differences in discharge questionnaire data**

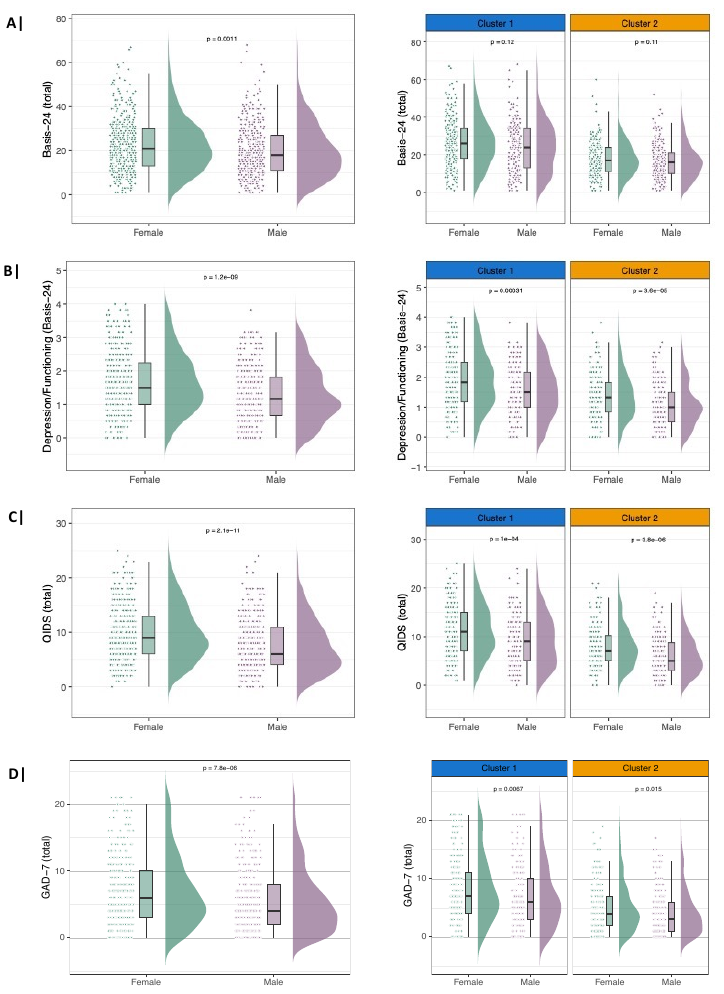

**Figure S5: Sex-related differences in discharge self-reported questionnaires.** Statistical comparison was performed using Kruskal-Wallis test.

**Section S7 Distributional differences in age and recruitment site: within–cohort and within-cluster**

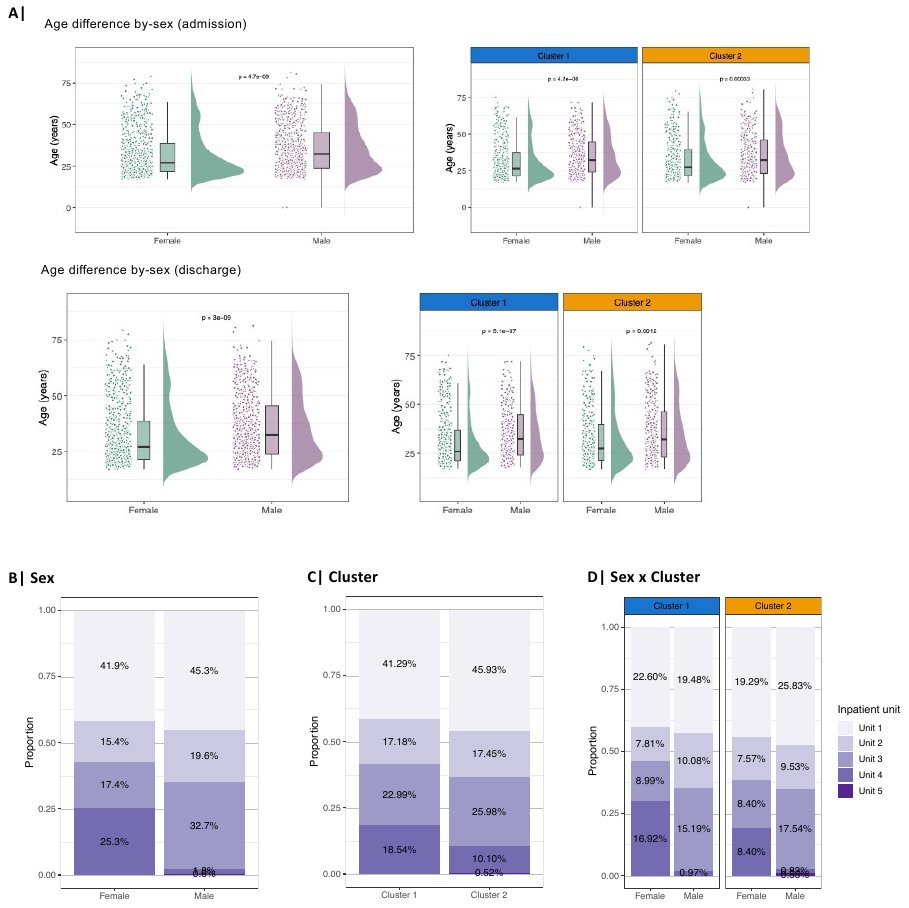

**Figure S6: Distribution differences in age and recruitment sites (inpatient units) within–cohort and within–cluster.** Statistical comparison was performed using Kruskal-Wallis test.

**Section S8 Between-cluster multi-granular functional connectivity differences**

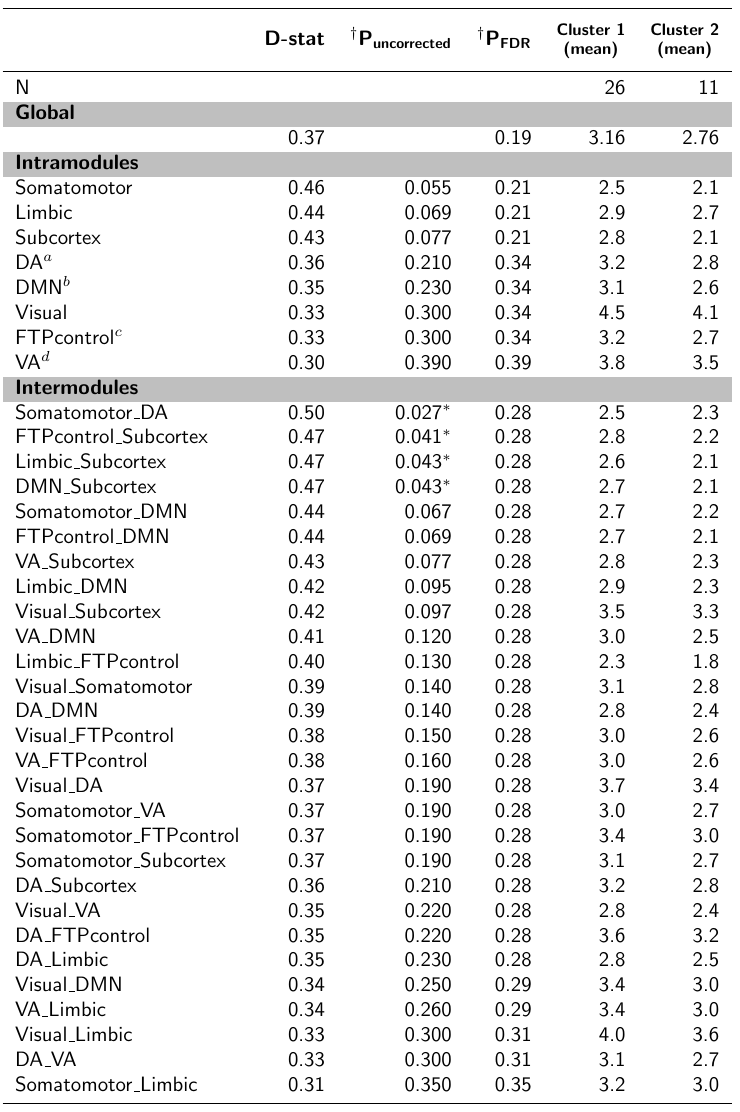

**Table S3: Between-cluster global and modular functional connectivity comparison.** Cortical functional network/modules are as defined by Yeo et al., (2011). *a*Dorsal attentional; *b*Default mode network; *c*Fronto-temporo-parietal control; *d*Ventral attentional; ∗P<0.05; ^†^statistical comparison performed using Kolmogorov-Smirnov test.

**Table S4: Between-cluster nodal functional connectivity comparison.** Nodes or ROIs are as defined by Glasser et al., (2016). Results are ordered by level of significance (Puncorrected).∗P<0.05; ^†^statistical comparison performed using Kolmogorov-Smirnov test.

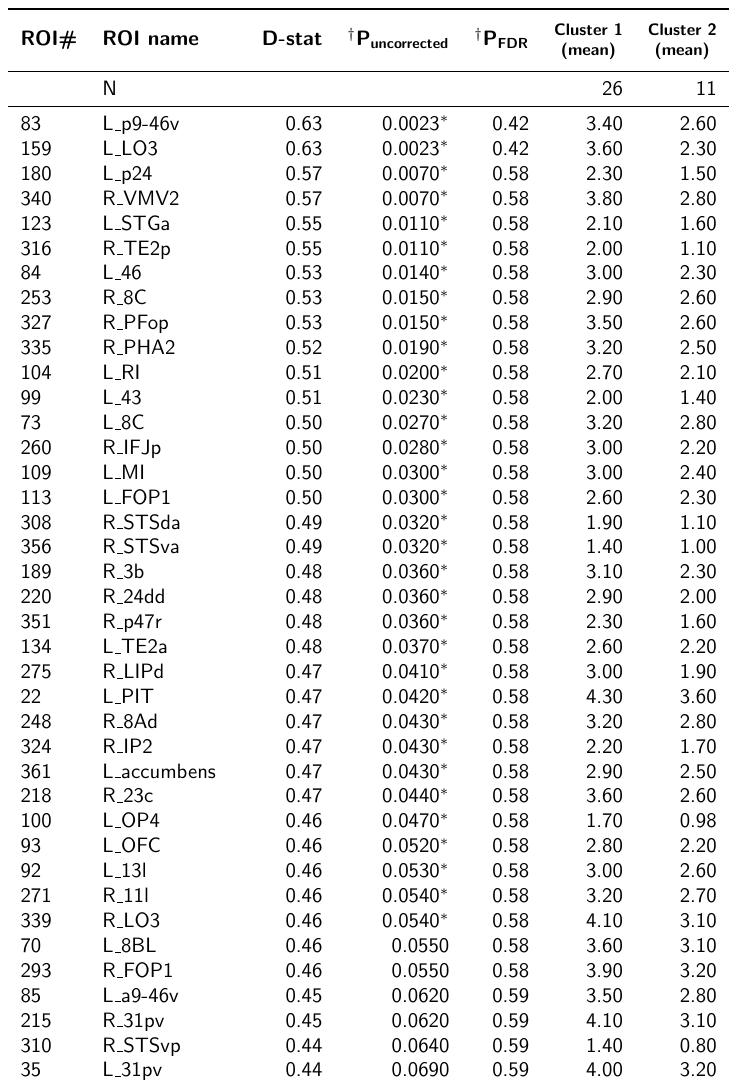

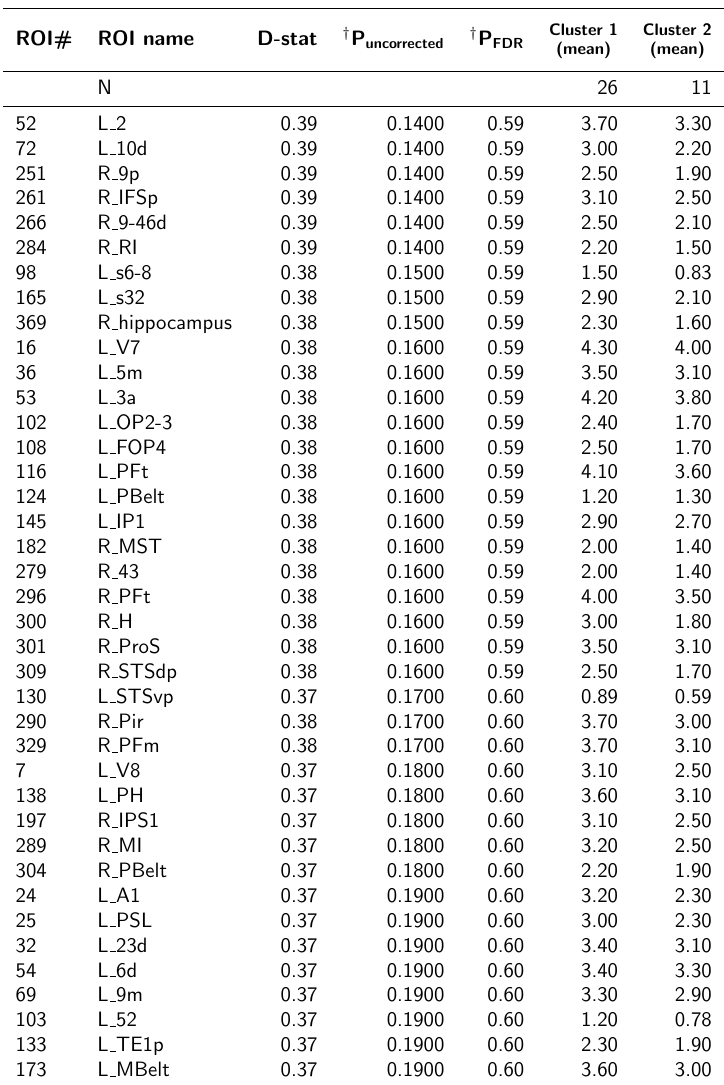

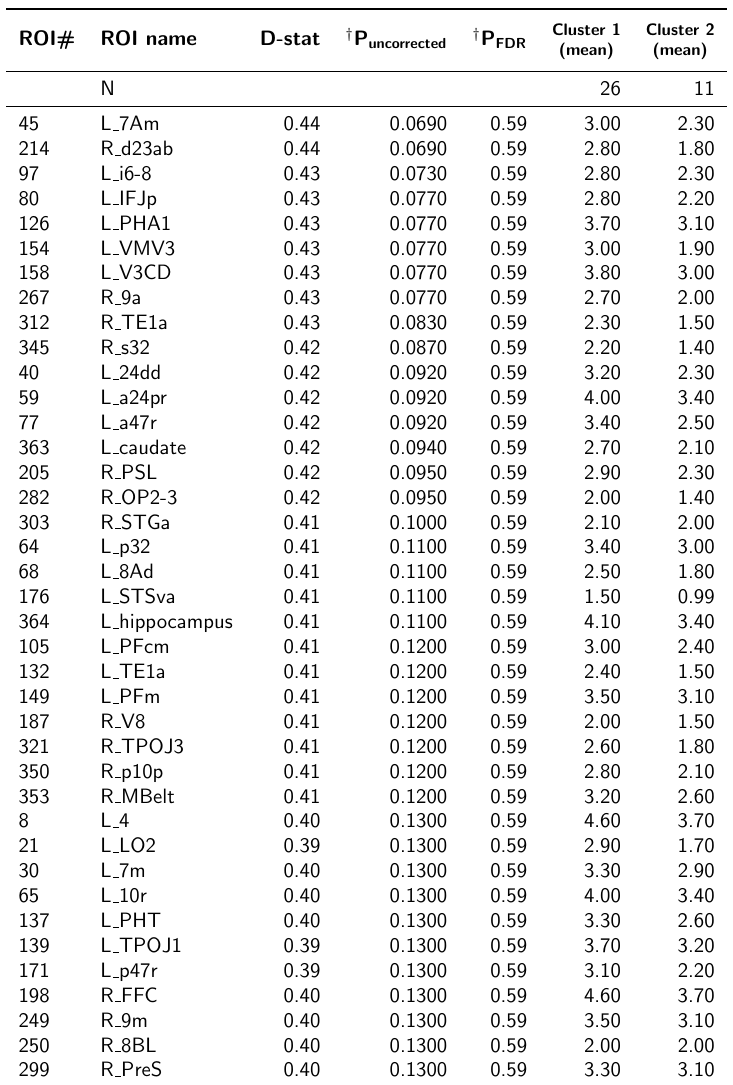

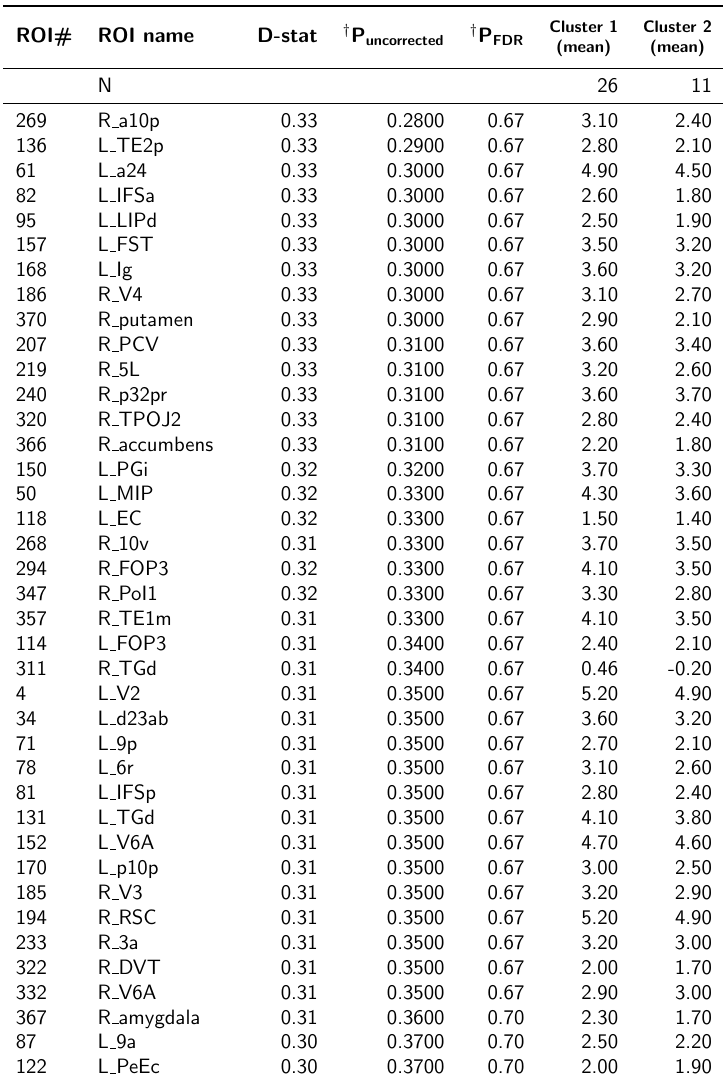

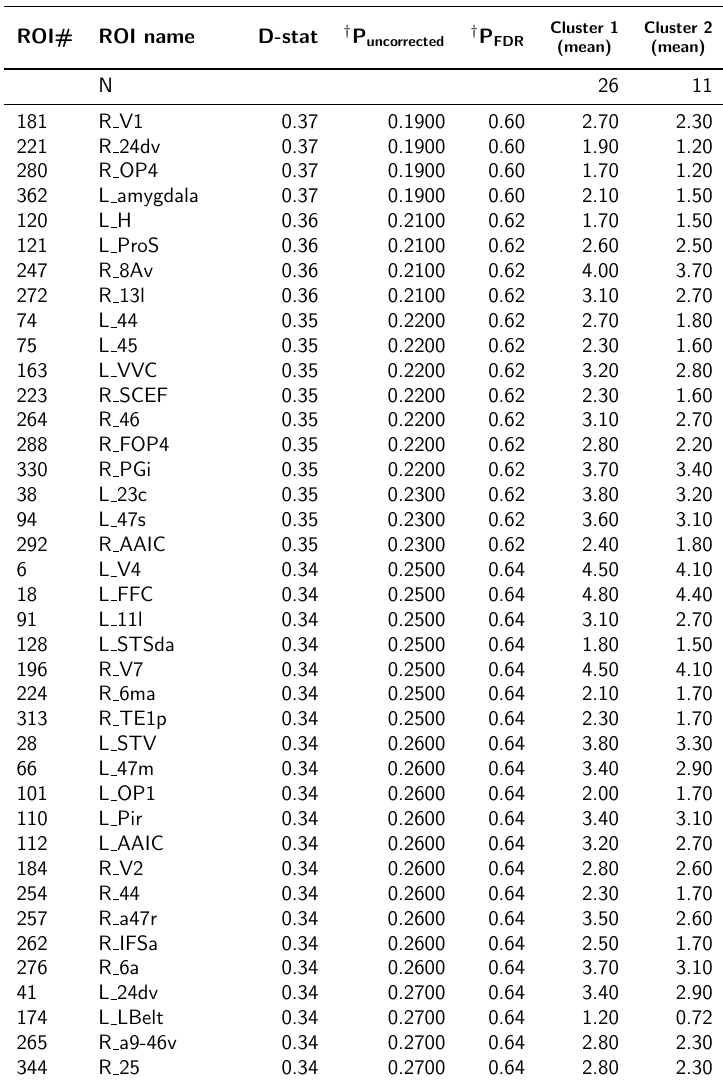

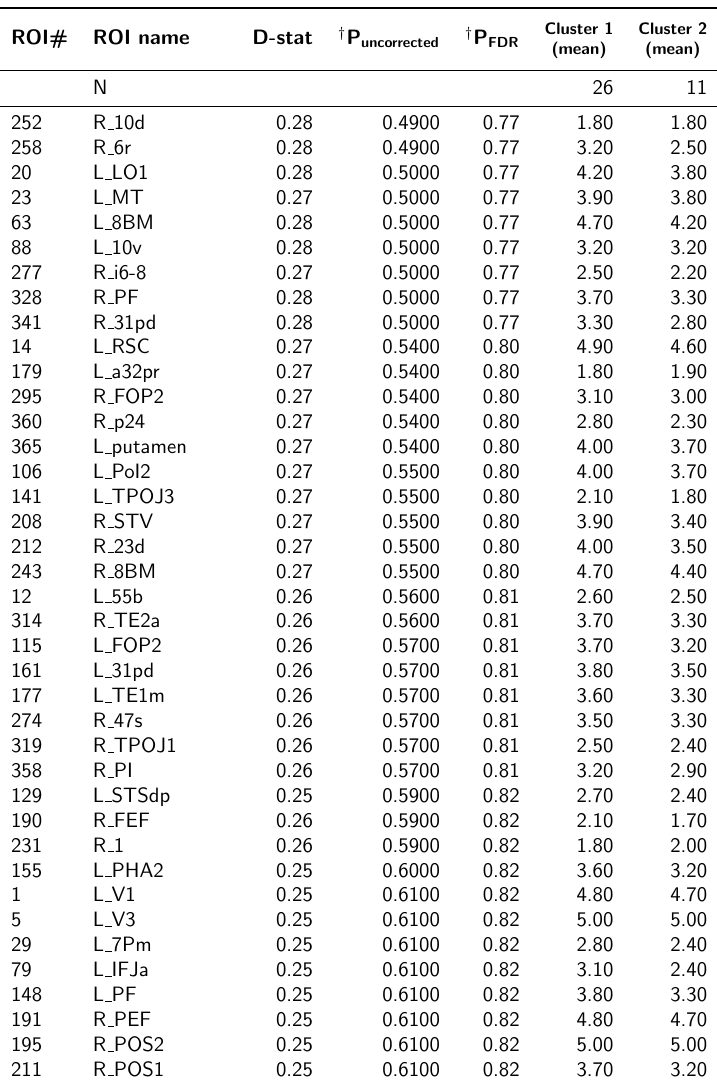

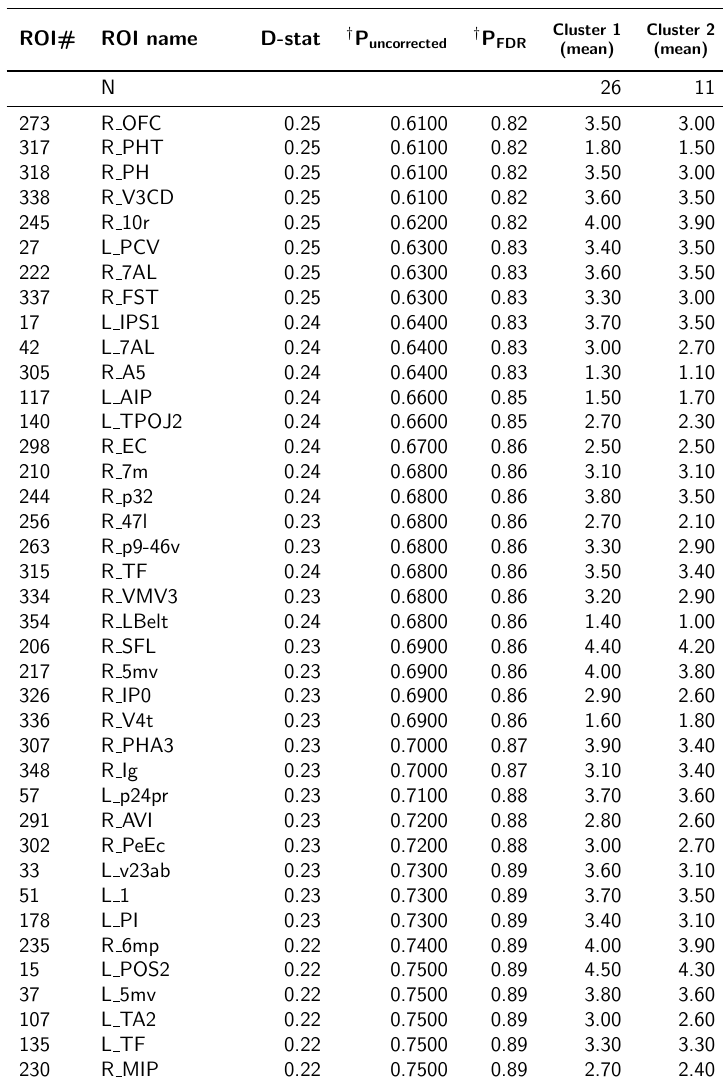

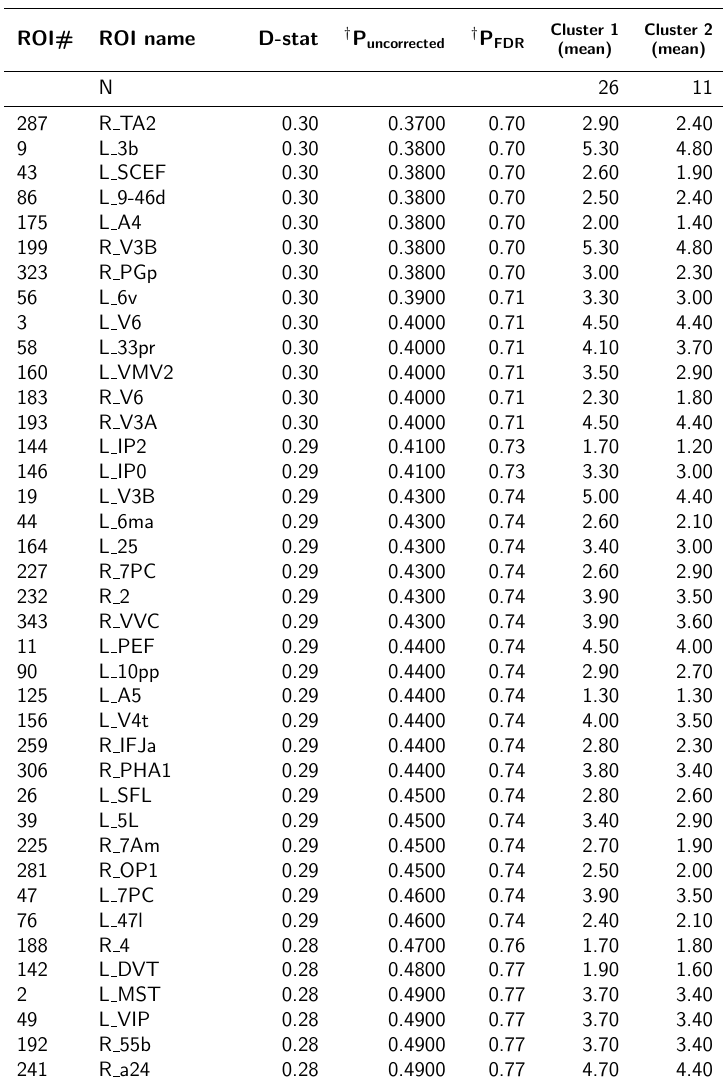

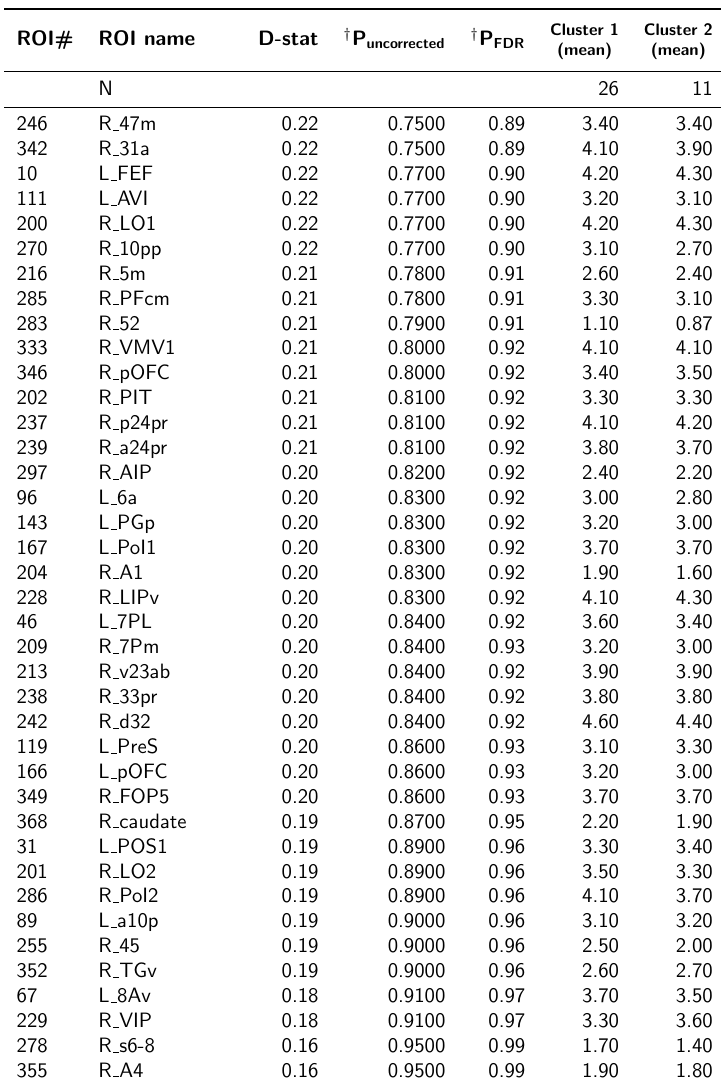

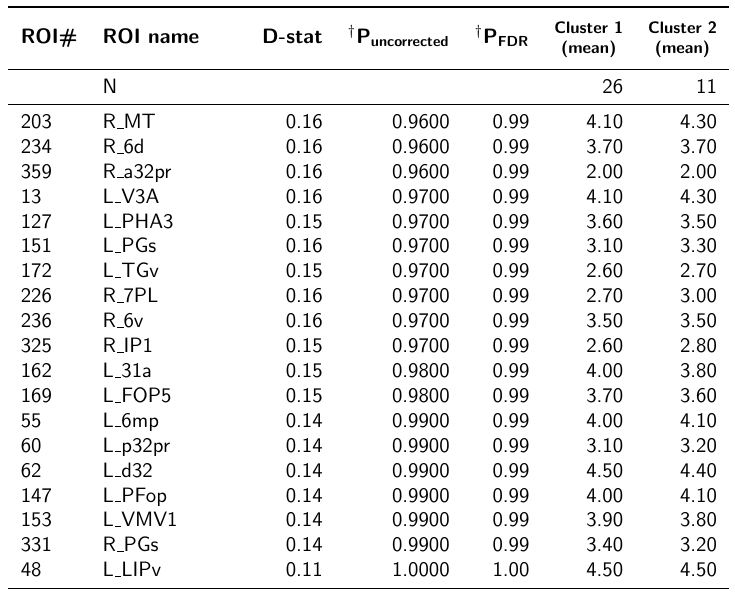
